# Poor sleep is robustly correlated with accelerated aging but the evidence for causation is mixed

**DOI:** 10.64898/2026.07.02.26357135

**Authors:** Ethan T. Whitman, Aric A. Prather, Julian Mutz, Louise Arseneault, David A.A. Baranger, Maxwell L. Elliott, Helen L. Fisher, David Ireland, Annchen R. Knodt, Christoph Kositzke, Yue Leng, Aaron Reuben, Karen Sugden, Benjamin S. Williams, J. Kathy Xie, Allison Yuan, Terrie E. Moffitt, Avshalom Caspi, Ahmad R. Hariri, Alzheimer’s Disease Neuroimaging Initiative

## Abstract

**SIGNIFICANCE STATEMENT:** Poor sleep increases with age and is correlated with increased risk of many different age-related diseases. This has led many to claim that poor sleep causes aging to accelerate. In our study, we tested this claim using data from over 64,000 people. We found clear evidence that poor sleep is correlated with faster aging during young, middle, and late adulthood. However, we found mixed evidence that poor sleep may cause aging to accelerate. Slowing aging by improving sleep warrants further research as causal claims are not consistently supported by the existing data.

Sleep gets worse with age and is correlated with risk for disease and mortality. The possibility that poor sleep causes aging to accelerate has prompted interest in improving sleep to slow aging and prevent disease. However, the existing evidence on the link between poor sleep and accelerated aging is unclear. Here, we tested for correlation and causation between poor sleep and accelerated aging using five independent datasets of adults (total N > 64,000). We found strong evidence for a correlation between poor sleep and fast aging that is consistent across young, middle, and late adulthood and across aging biomarkers derived from different tissues and modalities. We found that this correlation is robust to the influence of chronic disease burden, but not to the influence of shared genetic and early environmental factors among twins. Finally, we found mixed evidence for a causal influence of poor sleep on accelerated aging using Mendelian randomization. Our findings indicate that the correlation between poor sleep and accelerated aging is highly robust; however, the claim that poor sleep causes aging to accelerate is not consistently supported.

## INTRODUCTION

Sleep quality declines with age (1) and poor sleep is correlated with risk for many different age-related health conditions including cardiovascular disease, type 2 diabetes, obesity, and dementia (2–8). This has led to a widespread belief that poor sleep itself causes faster aging, as frequently claimed in popular media outlets (9–12) and on health blogs (13–16). This is an attractive possibility. Poor sleep is common and treatable (17, 18) and could therefore present an opportunity for early intervention to slow aging and prevent age-related diseases. However, the existing evidence on the link between sleep and aging has important limitations.

First, a major barrier to establishing a link between sleep and aging has been the measurement of aging itself. There are several studies documenting correlations between poor sleep and aging biomarkers (19–30). These biomarkers are difficult to compare with each other, making it challenging to consistently track associations between poor sleep and aging. These biomarkers are generally machine-learning models trained to predict chronological age or cross-sectional health or mortality risk from patterns of DNA methylation, brain structure, or other biological data. However, these biomarkers are each trained in a different dataset with different demographics and often a different training target phenotype entirely. As a result, these biomarkers cannot be easily compared to one another, making it challenging to assess the consistency of associations between poor sleep and aging biomarkers across these various studies.

In contrast, our group has developed multiple aging biomarkers from the same dataset and the same target phenotype but using different tissues and modalities. Specifically, we longitudinally tracked changes in 19 biomarkers of declining organ-system function from age 26–45 among members of the Dunedin Study, a longitudinal, population-representative study of a birth cohort of 1,037 people. We averaged the decline in the trajectories of these biomarkers into a specific measure that we called the Pace of Aging (31). We subsequently created two methods to estimate the Pace of Aging from different types of biological data. First, we created an algorithm that estimates the Pace of Aging from whole-blood DNA methylation called Dunedin Pace of Aging Calculated from the Epigenome, or DunedinPACE (32). Second, we created an algorithm to estimate Pace of Aging from structural brain MRI called Dunedin Pace of Aging Calculated from NeuroImaging, or DunedinPACNI (33). DunedinPACE and DunedinPACNI were trained in the same dataset with the same target phenotype (i.e., Pace of Aging) and have been extensively validated in independent datasets (32–35). Thus, DunedinPACE, and DunedinPACNI facilitate more interpretable comparison and could help test the consistency of associations between sleep and aging across different datasets.

Second, research on sleep and aging is largely unable to draw causal inferences due to observational designs (19–26, 28–30). One approach that can be used to strengthen evidence for a causal association is to rule out the influence of confounding variables. While some studies of poor sleep and aging have ruled out potential confounding from chronic disease (21, 24, 28, 29), there are many other potential confounders that are not measured or addressed. Comparing identical twins is an approach that can account for a large amount of confounding influence from shared genetic and early environmental factors (36, 37). Despite the strength of this approach, there is little research on the association between sleep and aging in twins (38), limiting conclusions on whether unmeasured genetic and/or early environmental factors confound the association between poor sleep and accelerated aging.

Further, there are mixed results from studies that explicitly test for a causal link between poor sleep and accelerated aging. There is experimental evidence in humans that acute sleep deprivation (>24 hours of wakefulness) may accelerate aging (27). However, this represents an extreme sleep disruption that may not generalize to the far milder levels of typical sleep disruption. Other studies have used Mendelian randomization (MR), a causal inference method that uses genetic variants as instrumental variables to account for measured and unmeasured confounding as well as reverse causality (39). MR studies have found mixed evidence for a causal influence of poor sleep on accelerated aging. Some sleep traits appear to have causal influence on some aging biomarkers (40–42), but most sleep traits and most aging biomarkers do not show evidence of a causal relationship (40–42). Furthermore, these studies use coarse measures of sleep, often with single self-report items and few response options. This can lead to selection of inappropriate genetic instruments and ultimately to misleading causal estimates (43).

Here, we sought to “stress-test” the association between poor sleep and fast aging. First, we used five independent datasets of adults (total N > 64,000) to test whether poor sleep was correlated with aging measured using both DunedinPACE and DunedinPACNI and whether this association was consistent in young, middle, and late adulthood. Second, we tested whether chronic disease burden, genetic makeup, or early environmental factors confound the associations between poor sleep and fast aging. Third, we explicitly tested for a causal relationship between poor sleep and fast aging by performing two-sample, bidirectional MR with genome-wide association study summary statistics from UK Biobank, 23andMe, FinnGen, and the Million Veterans Program.

## RESULTS

### Poor sleep is associated with faster aging across epochs of adulthood

We tested whether global sleep quality, as measured by the Pittsburgh Sleep Quality Index (PSQI; (44)), was correlated with the rate of aging in midlife. The PSQI generates a global sleep quality score based on self-reported problems such as difficulty falling asleep, frequency of waking during sleep, and discomfort during sleep (44).

First, we analyzed 1,207 participants from the MIDUS study with available DunedinPACE scores and PSQI data (mean (SD) age = 54.0 (12.6) years, age range = 26-86 years, 55.9% female). MIDUS participants reporting poorer global sleep quality were also aging faster when measured using DunedinPACE, the epigenetic measure of the Pace of Aging (*β*=0.23, *p*<0.001, 95% CI: 0.18-0.29; **Figure 1**). This analysis controlled for age and sex.

**Figure 1.**
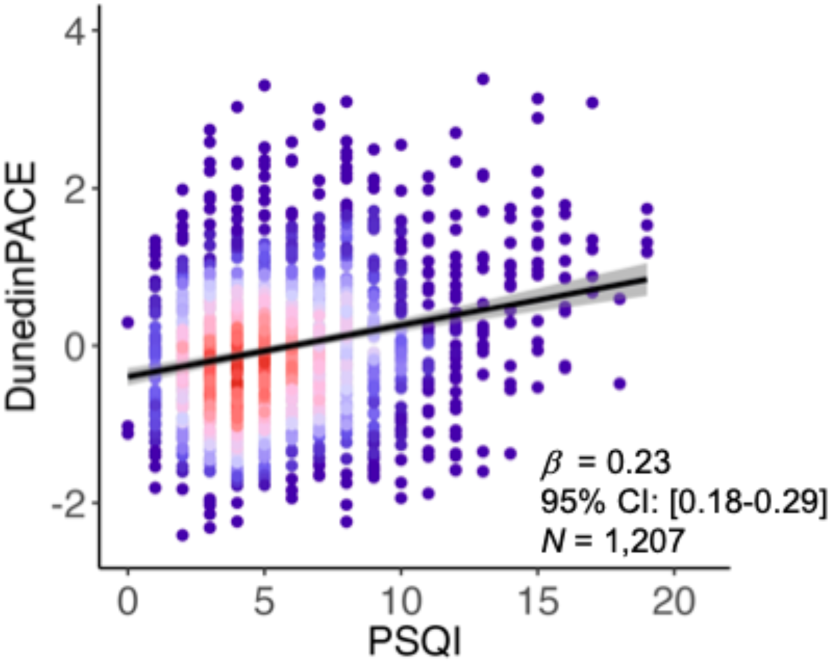
Poorer sleep quality is correlated with faster aging in midlife. Scatterplot of self-reported global sleep quality and DunedinPACE in MIDUS. Higher PSQI scores indicate poorer sleep quality. Each point represents an individual; warmer colors represent more individuals at that point. Abbreviations: CI = confidence interval, MIDUS = Midlife Development in the United States Study, PSQI = Pittsburgh Sleep Quality Index.

Previous research has shown that both abnormally short and long sleep duration are associated with poor health outcomes (45). Therefore, analyses next tested whether self-reported sleep duration was associated with faster aging in older adults from the UK Biobank. Aging was quantified in the UK Biobank using DunedinPACNI – the neuroimaging-based measure of the Pace of Aging. Among 59,844 participants with available data (mean age=65.0, SD age=7.74, age range=44-85, 53.2% female), we observed a U-shaped pattern such that those reporting shorter or longer sleep duration had faster DunedinPACNI scores (quadratic term *β*=0.36, *p*<0.001, 95% CI:0.31-0.41; vertex=7.1 hours; **Figure 2; Supplemental Figure S1**). When grouping UK Biobank participants according to the American Academy of Sleep Medicine designations of nightly sleep duration (i.e. short: <7 hours, normal: 7-9 hours, long: >9 hours) (46), participants with either short sleep or long sleep duration had faster DunedinPACNI than those with normal sleep duration (short: *β*=0.05, *p*<0.001, 95% CI:0.03-0.06; long: *β*=0.28, *p*<0.001, 95% CI:0.23-0.34). In addition, participants who endorsed “Usually” experiencing insomnia had slightly faster DunedinPACNI than those endorsing “Never/rarely” experiencing insomnia (*β*=0.02, *p*=0.02, 95% CI:0.003-0.04). These analyses covaried for age and sex.

**Figure 2.**
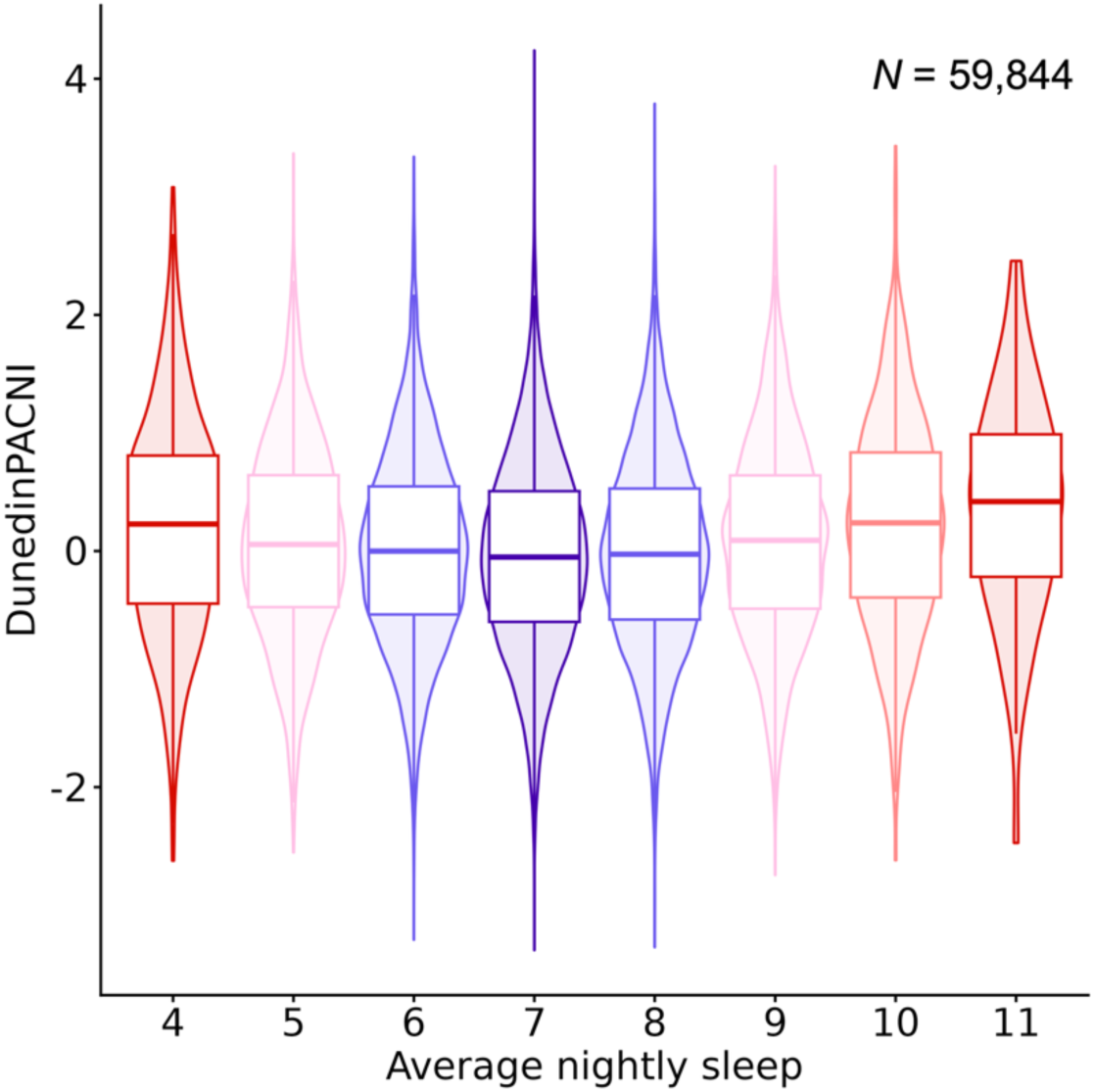
Midlife and older adults reporting either shorter or longer than average sleep duration were aging faster. Boxplot overlaid on violin plot of DunedinPACNI scores according to reported average hours of sleep per day. Cooler colors represent more normative sleep duration; warmer colors represent less normative sleep duration. Extreme outliers of average nightly sleep (<4 or >11 hours) were omitted to simplify visualization. A fitted quadratic regression model of sleep duration effect on DunedinPACNI with the full range of sleep duration is presented in **Supplemental Figure S1**.

Analyses next tested if a similar pattern was evident among even older adult participants in ADNI. Sleep quality was assessed through the Neuropsychiatric Inventory Questionnaire (NPIQ). The NPIQ is an informant-report questionnaire that includes the following question: “Does the patient awaken you during the night, rise too early in the morning, or take excessive naps during the day?”. If an informant responds “yes”, they then rate the severity of impairment as “Mild”, “Moderate”, or “Severe.” Aging was quantified using both DunedinPACNI (N = 759 individuals; 2,089 observations, mean (SD) age = 75.6 (6.7) years, 44% female) and DunedinPACE (N = 532 individuals; 809 observations, mean (SD) age = 75.4 (7.7) years, 44% female). Analyses revealed stepwise faster DunedinPACNI among participants reported to have “Mild” sleep impairment (*β*=0.26, *p*=0.002, 95% CI: 0.10-0.43), “Moderate” sleep impairment (*β*=0.35, *p*=0.002, 95% CI: 0.13-0.56), and “Severe” sleep impairment (*β*=0.87, *p*<0.001, 95% CI: 0.37-1.37) compared to those who were reported to have no sleep impairment (**Figure 3A**). A similar pattern was observed with DunedinPACE scores (“Mild” vs. none: *β*=0.22, *p*=0.06, 95% CI: −0.02-0.46; “Severe” vs. none: *β*=0.56, *p*=0.02, 95% CI: 0.09-1.02; **Figure 3B**), though not for participants reporting “Moderate” impairment (*β*=0.02, *p*=0.85, 95% CI: −0.20-0.24; **Figure 3B**). Note the reduced power in our analyses of DunedinPACE scores due to the smaller number of ADNI participants with available DNA methylation data. These analyses controlled for sex and age and included robust confidence intervals to account for individuals with multiple observations.

**Figure 3.**
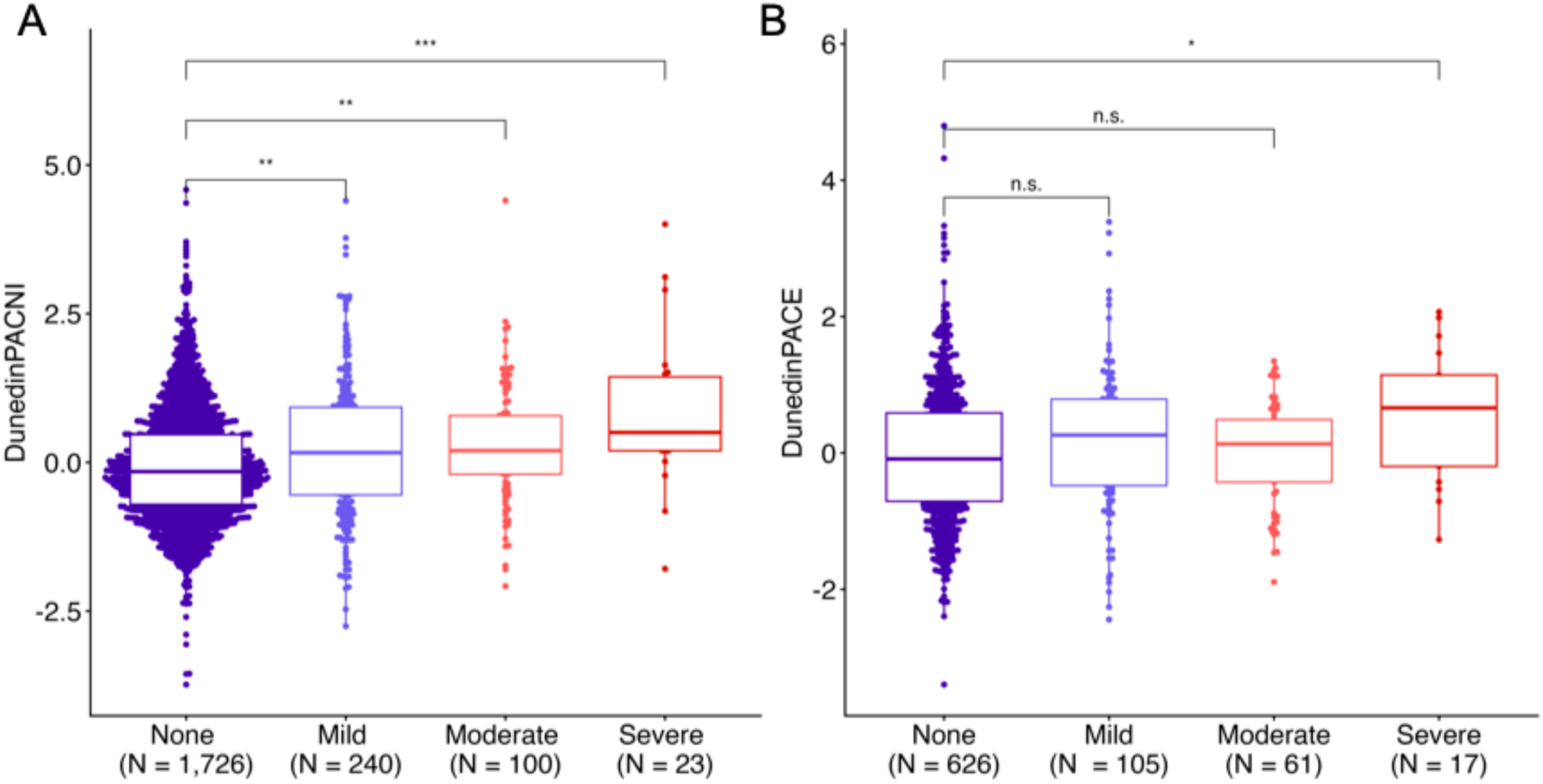
Older adults with greater informant-reported sleep impairment were aging faster. **A.** Boxplot of DunedinPACNI scores according to informant-reported sleep impairment among ADNI participants. **B.** Boxplot of DunedinPACE scores according to informant-reported sleep impairment among ADNI participants. Cooler colors represent less sleep impairment; warmer colors represent greater sleep impairment. Sample sizes presented on the x-axis reflect total number of observations, with some participants having multiple observations. Statistical significance was determined using robust standard errors to account for multiple observations per participant. Abbreviations: ADNI = Alzheimer’s Disease Neuroimaging Initiative, n.s. = not statistically significant, \**p* < .05, \*\**p* < .01, \*\*\**p* < .001.

We further tested whether the association between poor sleep quality and faster aging was present among younger adults, for whom chronic health problems have a much lower prevalence. Analyses were first performed on data from the Human Connectome Project (HCP), a sample of young adults free from psychiatric, neurologic, and cardiovascular disease (N = 1,113, mean age=28.8, SD age=3.7, age range=22-37, 54% female). Sleep quality was measured with the PSQI and aging was measured using DunedinPACNI. Consistent with findings in midlife and older adults, HCP participants reporting poorer global sleep quality had faster DunedinPACNI scores (*β*=0.08, *p*=0.004, 95% CI: 0.03-0.14; **Figure 4A**). This analysis covaried for age and sex.

**Figure 4.**
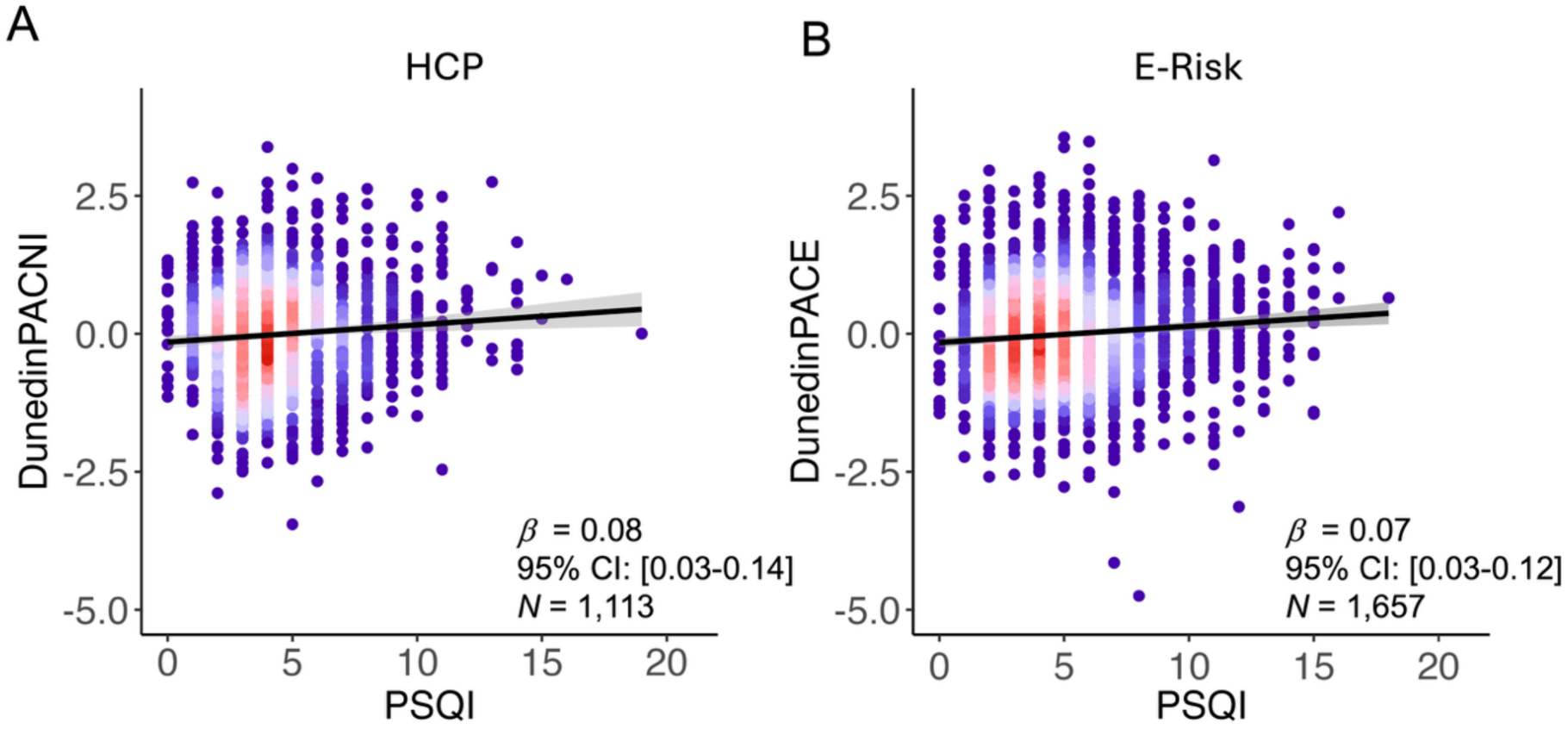
Poorer sleep quality is correlated with faster aging in young adulthood. **A.** Scatterplot of self-reported global sleep quality and DunedinPACNI in the HCP. **B.** Scatterplot of self-reported global sleep quality and DunedinPACE in E-Risk. Higher PSQI scores indicate poorer sleep quality. In each panel, each point represents an individual; warmer colors represent more individuals at that point. Abbreviations: CI = confidence interval, HCP = Human Connectome Project, PSQI = Pittsburgh Sleep Quality Index.

This finding was replicated using data from the Environmental Risk (E-Risk) Longitudinal Twin Study, a 1994-95 birth cohort of twins born in the United Kingdom (47), assessed when they were 18 years old (N = 1,657, 50% female). Sleep was measured with the PSQI and aging was measured with DunedinPACE. Again, E-Risk participants reporting poorer global sleep quality had faster DunedinPACE scores (*β*=0.07, *p*=0.002, 95% CI: 0.03-0.12; **Figure 4B**). This analysis covaried for sex and included a random effect for family.

### Associations between poor sleep and faster aging are not explained by chronic diseases

Poor sleep and faster aging are both associated with increased incidence of chronic diseases (2, 34). Therefore, we tested whether observed associations between sleep phenotypes and accelerated aging could be explained by the presence of chronic diseases. We repeated analyses in MIDUS and UK Biobank while controlling for indices of chronic disease (48) and in ADNI while controlling for mild cognitive impairment and dementia, which are the most common diagnoses in this dataset. Associations between poor sleep and faster aging survived these controls. In MIDUS, and UK Biobank, associations between poor sleep and faster aging were only modestly attenuated when controlling for chronic diseases (**Supplemental Tables S1-S2**). In ADNI, there was relatively greater attenuation of associations between poor sleep and DunedinPACNI, though not DunedinPACE, suggesting that brain diseases like dementia may have a stronger impact on the association between poor sleep and neuroimaging-based biomarkers of aging (**Supplemental Figure S2, Supplemental Table S3)**. Of note, the association between poor sleep quality and faster aging was present in HCP and E-Risk – two samples of young adults (E-Risk age = 18 years, HCP age range=22-37 years), well before the age of onset of most chronic diseases. This further supports the conclusion that the correlation between poor sleep and faster aging is not fully explained by chronic disease.

### Is the association between poor sleep and faster aging confounded by genetics or early environments?

Sleep quality and aging biomarkers are heritable (49, 50) and are affected by environmental factors shared by siblings growing up in the same family (e.g., parental health, early-life environment, socioeconomic background) (51–54). Sibling comparisons can be used to address genetic and environmental confounding factors that are shared between siblings, even when these factors are not measured directly. Dizygotic twins share the same household environment and, on average, 50% of their genetic material. Monozygotic twins share the same household environment and all of their genetic material. We tested whether correlations between poor sleep quality and faster aging were susceptible to confounding shared by siblings.

We analyzed 478 siblings and twins from the HCP dataset (N monozygotic = 240, N dizygotic = 238) and 1,466 twins from the E-Risk dataset (N monozygotic = 856, N dizygotic = 610). We first examined the within-family correlation of the PSQI and aging biomarkers in each dataset to verify sufficient within-family variation to warrant analysis. In HCP, we observed little within-family clustering of PSQI scores (monozygotic twins ICC = 0.14, dizygotic twins ICC = 0.17) and moderate within-family clustering of DunedinPACNI (monozygotic twins ICC = 0.65, dizygotic twins ICC = 0.37; **Supplemental Figure S3**). In E-Risk, we observed modest within-family clustering of PSQI scores (monozygotic ICC = 0.38, dizygotic = 0.07) and moderate within-family clustering of DunedinPACE (monozygotic ICC = 0.59, dizygotic ICC = 0.32; **Supplemental Figure S4**). This suggested there was sufficient within-family variation to examine within-family correlations between poor global sleep quality and faster aging.

Comparing pairs of monozygotic twins revealed that siblings with poorer sleep quality were not aging faster than their identical co-twins with better sleep quality (HCP: *β*=0.09, *p*=0.17, 95% CI: −0.04-0.22; E-Risk: *β*=0.02, *p*=0.48, 95% CI: −0.04-0.09; **Figure 5**). Likewise, comparing pairs of dizygotic twins revealed that siblings with poorer sleep quality were not aging faster than their co-twins with better sleep quality (HCP: *β*=0.12, *p*=0.07, 95% CI: −0.01-0.24; E-Risk: *β*=0.04, *p*=0.32, 95% CI: −0.04-0.12; **Figure 5**). Taken together, these findings provide weak evidence that the correlation between poor sleep and accelerated aging is independent from confounding genetic and environmental factors that are shared between siblings.

**Figure 5.**
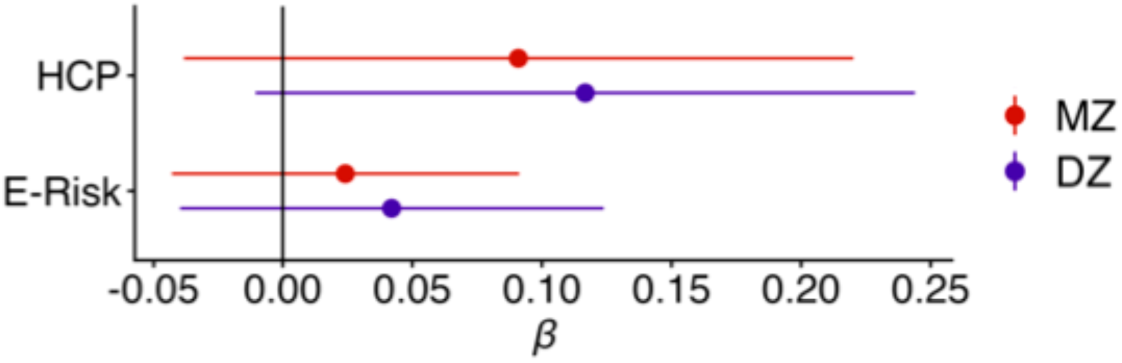
No evidence that the correlation between poor sleep and accelerated aging is independent from confounding genetic and environmental factors among twins. Forest plot of the within-family association between PSQI global sleep quality index and DunedinPACNI in HCP and with DunedinPACE in E-Risk. X-axis shows standardized effect size. Analyses in HCP covaried for age and sex. Analyses in E-Risk covaried for sex, but not age because all E-Risk participants were the same age. Abbreviations: DZ = dizygotic, HCP = Human Connectome Project, MZ = monozygotic, PSQI = Pittsburgh Sleep Quality Index.

### Evidence that clinical sleep disorders, though not self-reported insomnia, may cause faster aging

Given the robust correlation between sleep quality and the pace of aging, we tested whether self-reported insomnia or clinical sleep disorders cause faster aging using Mendelian randomization (MR). MR uses genetic variants such as single nucleotide polymorphisms (SNPs) as instrumental variables which can be used to draw causal inferences about relationships between phenotypes (39). We used summary statistics from four genome-wide association studies (GWAS) of sleep phenotypes and one GWAS of DunedinPACNI to perform two sample, bidirectional MR. Detailed results, including Manhattan plots, QQ plots, tests for heterogeneity, directional pleiotropy, and outlying SNPs, as well as the STROBE-MR and SLEEP-MR best practices reporting checklists can be found in **Supplemental Materials**.

We used summary statistics from two separate GWAS to identify genetic instruments for insomnia. Specifically, we first performed a GWAS of self-reported insomnia frequency in 244,436 unrelated UK Biobank participants of European ancestry. None of these participants had available neuroimaging data, thereby assuring no sample overlap between the insomnia and DunedinPACNI GWA studies. Second, we accessed summary statistics from a recent GWAS of self-reported insomnia performed in approximately 2 million participants from 23andMe, though only the top 10,000 SNPs were available for analysis (55). Finally, we performed a GWAS of DunedinPACNI scores in 65,868 UK Biobank participants of European ancestry. Further information on all three GWA studies can be found in **Supplemental Information S1-S2.**

We found no evidence that self-reported insomnia causes faster DunedinPACNI (UKB GWAS: 9 instrumental SNPs, minimum *F* statistic > 30, inverse-variance weighted *β*= −0.09, *p*=0.50, 95% CI: - 0.36-0.18; 23andMe GWAS: 105 instrumental SNPs, minimum *F* statistic > 51, inverse-variance weighted *β*=0.03, *p*=0.16, 95% CI: −0.01-0.08; **Figure 6A-B**). We performed several sensitivity analyses to examine the robustness of the MR results and potential bias due to horizontal pleiotropy, including leave-one-out, and MR-PRESSO outlier removal. These sensitivity analyses also provided limited evidence for a causal influence of self-reported insomnia on aging (**Supplemental Information S2**). Furthermore, sleep is influenced by highly pleiotropic genes (56, 57) and insomnia may lie downstream of other physiological changes that may also influence aging (e.g. obesity, hypoxia, inflammation). To test this possibility, we repeated these analyses while performing a biologically informed leave-many-out analysis wherein we excluded all SNPs that have been found to be associated with aging and/or neuroimaging phenotypes. This sensitivity analysis also provided limited evidence for a causal influence of self-reported insomnia on aging (**Supplemental Information S2**). We also did not find evidence for reverse causation (i.e. that faster DunedinPACNI scores causally influence self-reported insomnia, **Supplemental Information S2**).

**Figure 6.**
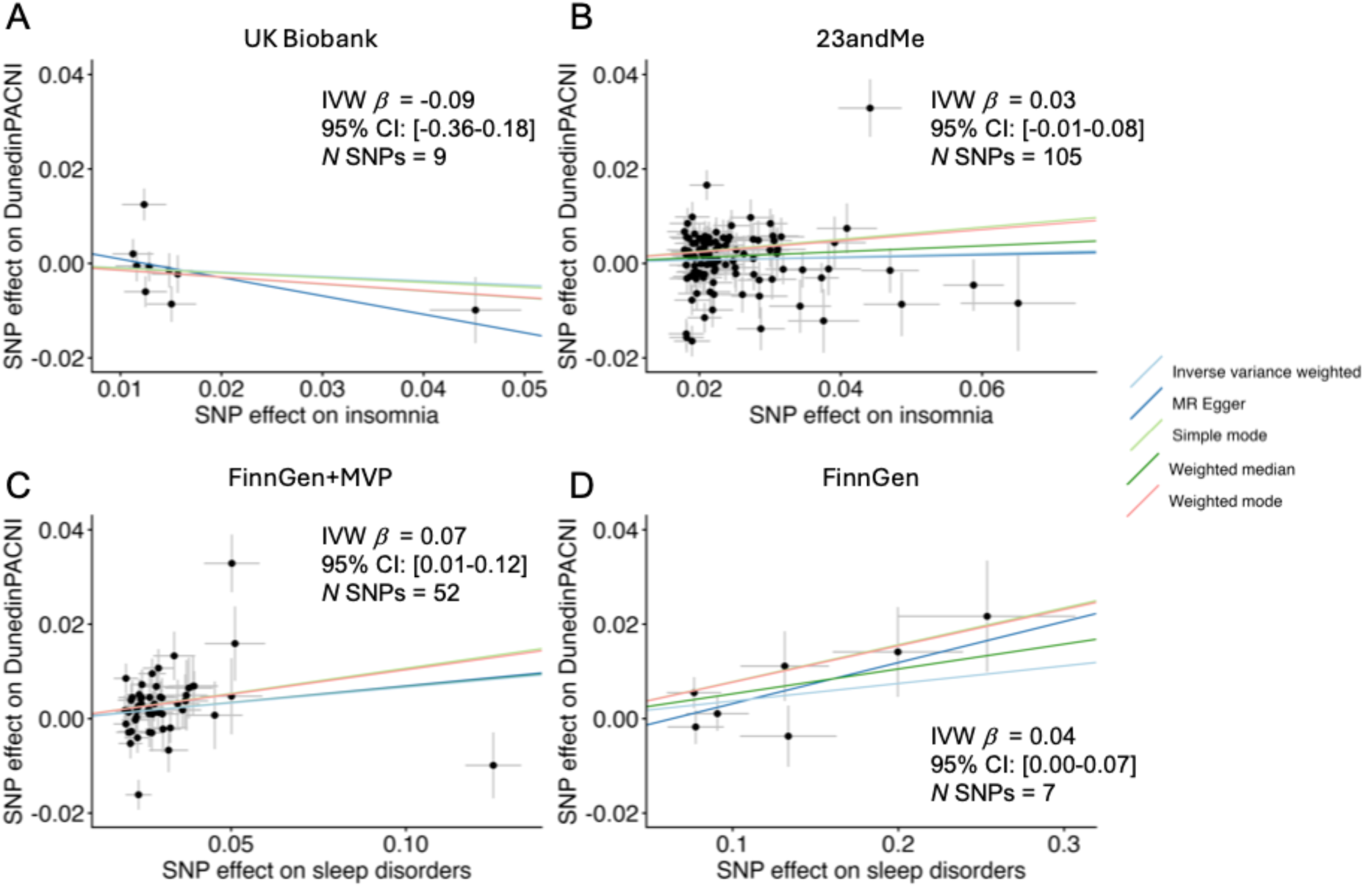
Mendelian randomization analyses suggest a causal influence of diagnosed sleep disorders, but not self-reported insomnia, on faster aging. **A.** SNP effects on self-reported insomnia in the UK Biobank non-imaging sample (x-axis) and on DunedinPACNI in the UK Biobank imaging sample (y-axis). **B.** SNP effects on self-reported insomnia in 23andMe (x-axis) and on DunedinPACNI in the UK Biobank imaging sample (y-axis). **C.** SNP effects on diagnosed sleep disorders in FinnGen and MVP (x-axis) and on DunedinPACNI in the UK Biobank imaging sample (y-axis). **D.** SNP effects on diagnosed nonorganic sleep disorders in FinnGen (x-axis) and on DunedinPACNI in the UK Biobank imaging sample (y-axis). Abbreviations: CI = confidence interval, IVW = inverse-variance weighted, MR Egger = Mendelian randomization Egger, SNP = single nucleotide polymorphism.

Of note, single-item self-reported sleep measures have been criticized for their poor agreement with objective sleep phenotypes, making them potentially inappropriate for MR (43). Therefore, we extended our analysis using a more objective measure of sleep disruption: diagnosis of a sleep disorder from medical records. Specifically, we used summary statistics from two additional GWAS to identify genetic instruments for diagnosed sleep disorders. First, we accessed summary statistics for any sleep disorder diagnosis (insomnia, hypersomnia, sleep-wake disorders, sleep terrors, nightmares, sleep disorders not otherwise specified, sleep apnea, narcolepsy and cataplexy, and other sleep disorders) from a meta-analysis of FinnGen and the Million Veterans Program (total N = 1,042,390). Second, we accessed summary statistics of a more restrictive GWAS of only non-organic sleep disorders in FinnGen (N = 370,640). Sleep disorders are considered non-organic when the disorder does not have any apparent physiological or neurological cause. The non-organic sleep disorder GWAS also excluded people with psychiatric or neurological disorders from the control group, as these disorders often include disrupted sleep as a symptom. More details on the sleep phenotypes used in these GWA studies, including specific ICD-10 codes, can be found in **Supplemental Information S3.**

In contrast to self-reported insomnia, we did find evidence that diagnosed sleep disorders cause faster DunedinPACNI (FinnGen and MVP GWAS: 52 instrumental SNPs, minimum *F* statistic > 29, inverse-variance weighted *β*=0.07, *p*=0.02, 95% CI: 0.01-0.12; FinnGen GWAS: 7 instrumental SNPs, minimum *F* statistic > 20, inverse-variance weighted *β*= 0.04, *p*=0.04, 95% CI: 0.00-0.07; **Figure 6C-D**). Alternative MR methods were consistent with the inverse-variance weighted estimator (**Figure 6C-D, Supplemental Information S3)** and MR-PRESSO adjustment for outlying SNPs strengthened evidence of causal influence of sleep disorders on DunedinPACNI (**Supplemental Information S3**). However, biologically informed leave-many-out analyses weakened the estimated causal effect of sleep disorders on DunedinPACNI when using genetic instruments from the FinnGen and MVP GWAS. Finally, we found limited evidence for reverse causation (i.e. that faster DunedinPACNI scores causally affect sleep disorders, **Supplemental Information S3**). Taken together, these results provide some evidence for a unidirectional causal relationship between diagnosed sleep disorders and faster aging.

## DISCUSSION

Our analyses of more than 64,000 people revealed a robust correlation between poor sleep quality and accelerated aging that is consistent across different biomarkers of aging and across young, middle, and late adulthood. However, there was mixed evidence for a causal relationship. Although the correlation was independent of the burden of chronic diseases, it was confounded by genetic and early environmental factors shared by monozygotic and dizygotic twins. Further, MR analyses did not suggest that self-reported insomnia causes faster aging, although they did provide evidence that medical-record diagnoses of sleep disorders, which included insomnia diagnoses, cause faster aging. These findings indicate that although the correlation between poor sleep and faster aging is robust, it remains uncertain whether poor sleep itself causes aging to accelerate.

Our study builds on previous reports of the association between poor sleep and accelerated aging (19, 20, 24–26, 29, 30, 58). Specifically, we found associations between sleep and the rate of aging that are consistent when aging is measured using DunedinPACE and DunedinPACNI. The fact that these methods of measuring aging yielded similar associations with poor sleep suggests that associations between sleep quality and aging biomarkers are not simply due to idiosyncratic aspects of measuring aging from the methylome or the brain but rather reflect robust associations between poor sleep and whole-body aging. Furthermore, we found that the association is apparent not only in midlife and older adults, but also in young adults. Most prior studies have focused on middle-aged and older adults (19, 20, 24–26, 29, 30, 58); our findings indicate that the link between poor sleep and faster aging is apparent in people as young as 18 years. This pattern is striking because many possible confounding factors that are common among older adults such as parenting, shift work, and chronic disease are much less prevalent among young adults. Future research should investigate whether the accelerated aging observed in young people represents very early effects of poor sleep or a signal of poorer system integrity (59) that also forecasts a predisposition to accelerated decline (60).

Analyses ruled out confounding from chronic disease, suggesting that chronic diseases do not fully explain the association between poor sleep and faster aging. We do note, however, that covarying for chronic diseases did attenuate the observed relationships, which were also weaker in young adults who did not have any chronic diseases. Therefore, it is possible that chronic diseases do explain some, though not all, of the association between poor sleep and fast aging. In contrast, our analyses of monozygotic and dizygotic twins suggest that the association between poor sleep and faster aging may arise due to shared early-environment and/or shared genetic factors. Indeed, sleep-linked genes are highly pleiotropic (56, 57), making it plausible that certain genetic variants that cause poor sleep also independently cause faster aging. An important caveat is that we were only able to perform our twin comparisons among relatively young adults (age range: 18-37 years).

MR analyses provided mixed evidence that poor sleep causes faster aging. MR analyses did not support a causal influence of self-reported insomnia on faster aging but did support a causal influence of sleep disorders as diagnosed from medical records on faster aging. There are several possible explanations for this discrepancy. First, it is possible that only sleep impairment severe enough to warrant clinical diagnosis causes faster aging and that the sleep disruption indexed by self-reported insomnia is too mild to exert a causal effect. This possibility is also supported by the weak observational association between self-reported insomnia frequency and DunedinPACNI in UK Biobank. Second, the insomnia phenotypes in UK Biobank and 23andMe were developed using very brief self-report items and show only moderate agreement with objective sleep phenotypes (61).

This may lead to poor genetic instruments which limit MR analyses (43). In contrast, the more objective sleep diagnosis phenotypes in FinnGen and the Million Veterans Program may have yielded better genetic instruments. Indeed, sleep disorder diagnosis is recommended over self-reported sleep in MR analyses (43). At present, however, it is unclear which of these possibilities explains our findings. We note that our mixed MR findings are consistent with the recent MR literature on sleep that has found limited and/or conflicting evidence for a causal effect of sleep on brain atrophy (62, 63), Alzheimer’s disease (64, 65), cancer (66), diabetes (67–69), and all-cause mortality (70). Future research, ideally experimental, should continue to examine whether sleep, or certain aspects of sleep, exert a causal influence on aging.

Our findings also build upon a recent report finding a remarkably consistent U-shaped relationship between reported sleep duration and older appearing organ function in midlife and older adults (30). We extend the findings of this report by showing that this association also appears between Pace of Aging biomarkers such as DunedinPACE and DunedinPACNI and that this pattern also exists among young adults. Further, this prior report performed mediation analyses to test potential mechanisms linking poor sleep, organ age gaps, and health outcomes. Here, using methods such as twin comparisons and MR, we more directly test for the possibility of a causal relationship of poor sleep on aging. Specifically, our analyses suggested that despite the clear correlation between sleep and aging phenotypes (30), the causal influence of sleep on aging remains ambiguous.

While we did not find clear evidence that poor sleep causes faster aging, this does not negate the numerous benefits of sleep. It is the overwhelming consensus of sleep researchers that sleep is critical for human health (46), sleep is physiologically necessary for survival in mammals (71–73), and sleep is nearly ubiquitous across the animal kingdom (74). Indeed, sleep has clearly documented causal benefits on cognition (75–77), mood (78), metabolic waste clearance in the brain (79), cardiovascular health (80), and immune system function (81–83). Furthermore, even though we did not detect a causal effect of poor sleep on aging, this does not preclude the possibility that sleep interventions could still have a positive effect on aging. Clinical trials of sleep interventions are needed to clarify potential geroprotective effects of sleep improvement, especially given the ambiguous evidence that poor sleep causes accelerated aging. Thus, while sleep plays a central role in health and could help to slow aging, we did not find clear evidence that poor sleep causes accelerated biological aging. We encourage careful delineation of these differences in future research (helpful recommendations are included in references 84 and 85).

There are several limitations to our analyses. First, participants in the UK Biobank, ADNI, the HCP, and E-Risk are primarily of European ancestry. Moreover, participants in the HCP and UK Biobank are healthier than the general population, and participants in the HCP, ADNI, and UK Biobank are more educated than the general population (86). Future research with datasets representing greater demographic and socioeconomic diversity will help establish the generalizability of associations between poor sleep and faster aging. Second, we largely focused on overall sleep quality, which combines many aspects of sleep including duration, falling asleep, staying asleep, and daytime wakefulness, among others. Likewise, our MR analyses tested the influence of all sleep disorder diagnoses, which includes many distinct disorders. This allowed us to investigate broad patterns of sleep disruption; however, future research should examine whether any of specific aspects of sleep behavior or any specific sleep disorders are uniquely related to aging. Additionally, our analyses were unable to test if specific components of sleep physiology (e.g., slow wave sleep, REM sleep) are related to faster aging. This is important given debates about how specific features of sleep physiology may impact risk for chronic age-related diseases such as Alzheimer’s disease (79, 87).

Third, we largely used self-reported measures of sleep, which can differ from objective measures (61). This problem is most apparent in UK Biobank, which uses only one question to measure certain sleep phenotypes. Of note, ordinal differences in objective sleep measures are generally consistent with even these coarse measures (61), which facilitates their use for correlational analysis. Likewise, we note that we primarily used the PSQI, which is a robust and well-validated measure of subjective sleep quality (44). Future studies utilizing objective measures of sleep will be helpful in further characterizing the associations between poor sleep quality and faster aging. Fourth, independent SNPs were selected using the European subset of the 1000 Genomes reference panel. A minority of participants in the FinnGen and MVP GWAS are from non-European ancestries (∼14%), and this could introduce bias in the selection of independent genetic instruments. We note that the GWAS of UK Biobank, 23andMe, and FinnGen were all restricted to participants of European ancestry. Fifth, we were unable to perform MR using GWAS of objectively measured sleep phenotypes. Although such GWAS have been performed in UK Biobank (88), there is substantial overlap among participants with objective sleep measures and those with neuroimaging, precluding two-sample MR. Future research should examine whether objectively measured sleep shows a clearer causal influence on aging.

Sleep is critical to health (46, 75–79), and public health messaging should clearly communicate this importance. However, public health messaging that sleeping badly accelerates biological aging is not clearly supported by the data (9–16). Such messaging ought to be restrained until the arrival of decisive evidence on the causal influence of sleep, ideally from clinical trials.

## MATERIALS AND METHODS

Analyses were conducted on data collected through MIDUS, UK Biobank, ADNI, the HCP, and E-Risk Study. Details for each study and dataset are described below. All analyses and code were checked for accuracy by an independent analyst.

### Data Sources

#### Midlife in the United States

The Midlife in the United States (MIDUS) study is a nationwide longitudinal study of noninstitutionalized, English-speaking adults in the United States (89, 90). We analyzed participants from the MIDUS core sample and from the MIDUS Refresher sample. The MIDUS core sample were adults aged 25-74 in 1995-1996 (N = 7,108). The MIDUS Refresher sample was a probability sample aged 24-74 in 2011-2014 who underwent the same comprehensive assessments as the MIDUS core sample. Specifically, we analyzed data from the second assessment of the MIDUS core sample and the first assessment of the MIDUS Refresher sample. The MIDUS study was approved by the institutional review boards of the University of Wisconsin, Madison and the Pennsylvania State University. All MIDUS participants provided written informed consent. MIDUS sample demographic information can be found in **Supplemental Table S4.** Detailed inclusion criteria are presented in **Supplemental Figure S5**.

### DNA Methylation

The Illumina Infinium MethylationEPIC microarray was used to measure DNA methylation from whole blood (91). Details of the MIDUS DNA methylation data have been published elsewhere (92). Detailed inclusion and exclusion information is presented in **Supplemental Figure S5**.

### Sleep Quality

The Pittsburgh Sleep Quality Index (PSQI) was completed during MIDUS assessment II and MIDUS Refresher assessment I. The PSQI is a 19-item self-report measure of seven sleep categories: subjective sleep quality, sleep latency, sleep duration, habitual sleep efficiency, sleep disturbances, use of sleeping medication, and daytime dysfunction (44). The PSQI yields a measure of global sleep quality with scores ranging from 0-21, with higher scores indicating lower global sleep quality. PSQI global sleep quality scores at age 45 were used in analyses.

### Current Health Conditions

Current health conditions were assessed according to self-reported diagnoses of health conditions at the time of assessment. The conditions included were stroke, Parkinson’s disease, cardiovascular disease, cancer, asthma/bronchitis/emphysema, other lung problems, joint/bone disease, AIDS/HIV, diabetes/high blood sugar, and ulcer. We calculated a summary score of overall chronic disease burden with a count of the total number of endorsed conditions.

#### UK Biobank

The UK Biobank is a United Kingdom population-based prospective study of 502,486 participants between the ages of 37 and 73 at baseline assessment (93). UK Biobank was approved by the North West Multi-Centre for Research Ethics Committee. All UK Biobank participants provided written informed consent. UK Biobank sample demographic information can be found in **Supplemental Tables S5-S6**.

### Neuroimaging

MRI data were collected using three identical 3T Siemens Skyra scanners with a 32-channel Siemens head coil (94). T1-weighted images were obtained using a 3D MP-RAGE with the following parameters: TR=2000 ms; TI=880 ms; 208 sagittal slices, matrix=256×256; slice thickness=1 mm with no gap; and total scan time=4 min and 52 s. Raw T1-weighted images were processed using cross-sectional FreeSurfer version 6.0 (94). All brain measures used in UK Biobank cross-sectional analyses presented herein were derived from this FreeSurfer pipeline. We excluded UK Biobank participants with very low signal-to-noise ratio and highly unusual summary morphometrics indicative of low-quality reconstruction. Detailed inclusion and exclusion information is presented in **Supplemental Figure S6**.

### Sleep Quality

Contemporaneous sleep was measured using two self-report items. First, sleep duration was measured as the reported average number of hours slept per 24 hours, including naps (Data Field 1160). Responses <1 hour or >23 hours were rejected. Participants were asked to confirm responses <3 hours or >12 hours. Second, insomnia was measured using the question, “Do you have trouble falling asleep at night or do you wake up in the middle of the night?”, to which participants selected “Never/rarely,” “Sometimes,” “Usually,” or “Prefer not to answer” (Data Field 1200). Participants completed these sleep self-report items during the same visit as MRI scanning.

### Current Health Conditions

We derived the Charlson Comorbidity Index (48) to measure participants’ current health status. The Charlson Comorbidity Index is the weighted sum of comorbid chronic diseases with more severe diagnoses assigned greater weight. Diagnoses were assessed using lifetime hospital inpatient ICD-10 diagnostic codes (Data field 41270), as described previously (95). A full list of ICD-10 codes used in calculating the Charlson Comorbidity Index is presented in **Supplemental Table S7**.

### Genetics

We used imputed SNP data from UK Biobank participants. The details of collection and processing of these data have been reported elsewhere (96). Detailed inclusion and exclusion information for UK Biobank samples used in GWAS and MR analyses is presented in **Supplemental Figure S6**. Detailed QC of SNPs are presented in **Supplemental Information S1-S3.**

#### Alzheimer’s Disease Neuroimaging Initiative

Data used in the preparation of this article were obtained from the Alzheimer’s Disease Neuroimaging Initiative (ADNI) database (adni.loni.usc.edu). The ADNI was launched in 2003 as a public-private partnership, led by Principal Investigator Michael W. Weiner, MD. The primary goal of ADNI has been to test whether serial magnetic resonance imaging (MRI), positron emission tomography (PET), other biological markers, and clinical and neuropsychological assessment can be combined to measure the progression of mild cognitive impairment (MCI) and early Alzheimer’s disease (AD) (97). Cognitive and diagnostic data were downloaded on June 12^th^, 2022. MRI data curated from the Alzheimer’s Disease Sequencing Project collection were downloaded on December 7^th^, 2023. ADNI was approved by the Institutional Review Boards of all the participating institutions. All ADNI participants provided written informed consent. ADNI sample demographic information can be found in **Supplemental Table S8**.

### Neuroimaging

T1-weighted scans were collected using either 1.5T or 3T scanners. MRI acquisition parameters varied across ADNI sites and waves; however, the targets for acquisition were isotropic 1mm^3^ voxels (98). Raw T1-weighted images were processed using longitudinal FreeSurfer version 6.0. Images were excluded for low quality if they did not have a QC rating of ‘Pass’ from ADNI investigators or if segmentation failed visual inspection. Images were also excluded if participants were missing demographic data such as age, sex, or diagnosis. Detailed inclusion and exclusion information for the ADNI imaging sample is presented in **Supplemental Figure S7.**

### DNA Methylation

The Illumina Infinium MethylationEPIC BeadChip Array was used to measure DNA methylation from whole blood (99, 100). Detailed inclusion and exclusion information for the ADNI DNA methylation sample is presented in **Supplemental Figure S8.**

### Sleep Quality

The Neuropsychiatric Inventory Questionnaire (NPIQ) was used to measure sleep quality. The NPIQ is an informant report questionnaire that includes the following question regarding sleep quality: “Does the patient awaken you during the night, rise too early in the morning, or take excessive naps during the day?” The informant then selects “yes” or “no.” If the informant selects “yes,” they then indicate the severity of this problem as “Mild”, “Moderate”, or “Severe”. Informants were friends or family members who knew participants well, could answer questions about their day-to-day functioning, and accompanied them to the study visit. Sleep quality observations were paired with MRI scans that had occurred within one month. Sleep quality observations were paired with DNAm blood draws that had occurred within six months due to the relatively smaller number of DNAm observations in ADNI.

### Cognitive Status

Participants were classified into cognitively normal (CN), mild cognitive impairment (MCI), or dementia groups by ADNI study physicians based on subjective memory complaints, multiple neurocognitive and behavioral assessment scores, and level of impairment in activities of daily living (for more detailed criteria see https://adni.loni.usc.edu/data-samples/adni-data/study-cohort-information/). Each individual MRI scan was categorized according to the most temporally proximate cognitive diagnosis received by that participant.

#### Human Connectome Project

The HCP is a publicly available dataset that includes 1,206 participants with extensive MRI and behavioral data (101). HCP data access is managed by the WU-Minn HCP consortium. All participants provided written informed consent. HCP participants were excluded if they reported history of premature birth, head injury, as well as diagnosis, hospitalization, or significant treatment for any psychiatric disorder, substance abuse, neurological, or cardiovascular disease, as has been described in detail elsewhere (101). HCP sample demographic information can be found in **Supplemental Table S9** and HCP inclusion and exclusion criteria for analyses in this manuscript is presented in **Supplemental Figure S9.**

### Neuroimaging

Structural MRI data were analyzed using the HCP minimal preprocessing pipeline (102). Briefly, T1-weighted images were processed using a custom FreeSurfer recon-all pipeline that is optimized for structural MRI with a higher resolution than 1 mm isotropic. Details about MRI data acquisition have been described elsewhere (102).

### Sleep Quality

The PSQI was completed by participants at the time of scanning.

#### Environmental Risk Longitudinal Twin Study

The Environmental Risk (E-Risk) Longitudinal Twin Study is a cohort of 2,232 twins born in the United Kingdom followed from birth to age 18 years. Full details on the sample have been reported previously (47). E-Risk Study sample demographic information can be found in **Supplemental Table S10.**

Briefly, the E-Risk Study sample was constructed in 1999–2000, when 1,116 families (93% of those eligible) with same-sex 5-year-old twins participated in home-visit assessments. This sample comprised 56% monozygotic (MZ) and 44% dizygotic (DZ) twin pairs. The study sample represents the full range of socioeconomic conditions in Great Britain, as reflected in the families’ distribution on a neighborhood-level socioeconomic index (103). Follow-up assessments were conducted when the participants were aged 5, 7, 10, 12, and 18 years. There were 2,066 children who participated in the E-Risk Study assessments at age 18, and the proportions of MZ (56%) and male same-sex (47%) twins were almost identical to those found in the original sample at age 5. The Joint South London and Maudsley, and the Institute of Psychiatry Research Ethics Committee approved each phase of the study. Parents gave informed consent, and twins gave assent between 5 and 12 years and then informed consent at age 18.

### DNA Methylation

The Illumina 450k array was used to measure DNA methylation from whole blood collected at age 18 (32). Detailed inclusion and exclusion information for the E-Risk Study DNA methylation sample is presented in **Supplemental Figure S10.**

### Sleep Quality

The PSQI was completed by participants during the age 18 assessment.

### Measures of Aging

DunedinPACE and DunedinPACNI are both versions of the same measure. We use DunedinPACE when we report from a study that lacks repeated biomarkers to build Pace of Aging but has whole-blood DNA methylation, and we use DunedinPACNI when we report from a study that lacks repeated biomarkers to build Pace of Aging but has brain MRI. Details of each measure are elaborated below.

### DunedinPACE

We derived DunedinPACE from DNA methylation data using the publicly available algorithm (https://github.com/danbelsky/DunedinPACE). Briefly, elastic net regression was used to predict Pace of Aging from a set of 173 CpG sites derived from whole blood from Dunedin Study members (Belsky et al., 2022). We then applied regression weights to the methylation scores at these CpG sites in new datasets to calculate DunedinPACE scores. Within each dataset, DunedinPACE values were standardized to mean=0, standard deviation=1, as done in previous studies (32, 99).

### DunedinPACNI

We derived DunedinPACNI from T1-weighted MRI scans of the brain using the publicly available algorithm (https://github.com/etw11/DunedinPACNI). Briefly, elastic net regression was used to predict the Pace of Aging from a set of 315 structural brain measures extracted from T1-weighted images. These measures included regional cortical thickness and surface area, regional cortical gray matter volume and gray-white matter signal intensity ratio, subcortical gray matter volumes, ventricular volumes, and bilateral volume of white matter hypointensities. We then applied regression weights to these structural MRI measures in new datasets to calculate DunedinPACNI scores. Within each dataset, DunedinPACNI values were standardized to mean=0, standard deviation=1, as done in previous studies (33, 104).

### Statistical Analyses

Testing cross-sectional associations between sleep quality and aging.

We first tested for cross-sectional associations between poor sleep quality and aging in MIDUS, UK Biobank, ADNI, HCP, and E-Risk Study. Specifically, in MIDUS, we tested for a linear association between PSQI global sleep quality scores and DunedinPACE. In UK Biobank, we first tested for a quadratic association between contemporaneously reported sleep duration and DunedinPACNI scores (N=59,844) because both abnormally short and long sleep duration have been associated with poor health outcomes (45). We next tested if DunedinPACNI was faster among participants who contemporaneously reported that they “Sometimes” or “Usually” experienced insomnia. In ADNI, we tested for stepwise faster DunedinPACE (N=809 observations, 532 individuals) and DunedinPACNI (N=2,089 observations, 759 individuals) amongst participants with greater informant-reported severity of sleep problems. In the HCP, we conducted a linear regression between PSQI global sleep quality scores and DunedinPACNI (N=1,113). In E-Risk, we conducted a linear regression between PSQI global sleep quality scores and DunedinPACE (N = 1,657) while including a random effect for family. All cross-sectional analyses controlled for sex. Analyses in MIDUS, UK Biobank, ADNI, and the HCP also controlled for age, though this was not necessary in the E-Risk Study because all participants were the same age.

### Testing whether chronic disease burden accounts for associations between sleep quality and aging

We tested whether associations between poor sleep quality and faster aging could be explained by overall poor health by repeating all analyses while controlling for the presence of chronic diseases.

Specifically, chronic disease burden was measured in MIDUS as the count of endorsed chronic health conditions, in UK Biobank using the Charlson Comorbidity Index (48), and in ADNI using cognitive status diagnosis. Analyses in all datasets controlled for age and sex.

### Testing whether genetics or home environment explain the association between sleep quality and aging

We tested whether associations between poor sleep quality and faster aging exist within monozygotic and dizygotic twins. To do so, we first analyzed 478 siblings from 239 families from the HCP dataset. We calculated within-family differences in PSQI global sleep quality scores and DunedinPACNI by finding the difference between each person’s score and the average for their family. Next, we divided the sample into monozygotic (N = 240) and dizygotic twins (N = 238). We then tested for linear relationships between within-family differences in PSQI global sleep quality scores and within-family differences in DunedinPACNI while covarying for age and sex within each zygosity group.

We conducted parallel analyses using data from 1,466 twins from the E-Risk Study where both twins had available PSQI and DunedinPACE scores. We again estimated within-family differences in PSQI global sleep quality scores and DunedinPACE. We calculated these differences by finding the difference between siblings, as there were only two participants per family. Next, we divided the sample into monozygotic twins (N = 856) and dizygotic twins (N = 610). We tested for linear relationships between within-family differences in PSQI global sleep quality scores and within-family differences in DunedinPACE while covarying for sex within each zygosity group. It was not necessary to covary for age because all E-Risk Study participants were 18 years old.

### Testing whether sleep phenotypes cause faster aging

Mendelian randomization (MR) is an instrumental variable method that treats genetic variants, such as single-nucleotide polymorphisms (SNPs), as genetic instruments and can be used for causal inference (39). MR is based on three core assumptions: (i) relevance, the genetic instrument must be strongly correlated with the exposure, (ii) independence, the genetic instrument must not share any common causes with the outcome, and (iii) exclusion, the genetic instrument must only affect the outcome through the exposure (39). All three assumptions must be met for valid causal inference, though only the relevance assumption can be empirically verified.

We conducted four different two-sample MR analyses to test the causal influence of sleep phenotypes on DunedinPACNI scores. First, we performed a genome-wide association study (GWAS) of self-reported insomnia frequency (Data Field 1200) in 244,436 unrelated UK Biobank participants of European ancestry who did not have neuroimaging data (to ensure no overlap between samples). Participants were asked, “Do you have trouble falling asleep at night or do you wake up in the middle of the night?” on a touchscreen. Insomnia frequency was treated as a quantitative variable from 0-2 with responses of “Never/rarely” coded as 0, “Sometimes” coded as 1, and “Usually” coded as 2. The GWAS was performed using REGENIE (105) for all SNPs that passed quality control metrics (INFO > 0.8, minor allele count > 5, minor allele frequency > 1%, missing data < 10%, and Hardy-Weinberg equilibrium *p* < 1e-6). GWAS covariates included age, genetic sex, batch, and genetic principal components 1-10. We did not observe notable genomic inflation (λ_1000_ = 1.000029). Further details of this GWAS can be found in **Supplemental Information S1.** Next, we accessed summary statistics from three independent GWAS: (i) the top 10,000 SNPs associated with reported insomnia from 1,978,022 participants from 23andMe (55), (ii) full, meta-analyzed summary statistics of a GWAS of sleep disorders performed in FinnGen and the Million Veteran Program (MVP; N = 1,042,390) and (iii) full summary statistics of a GWAS of non-organic sleep disorders performed in FinnGen (N = 370,640). Summary statistics for the FinnGen and MVP meta-analysis and the FinnGen GWAS are available at https://r12.finngen.fi.

Finally, we performed a GWAS of DunedinPACNI using 65,868 UK Biobank participants of European ancestry. We found that approximately 4% of this sample showed some degree of cryptic relatedness. We elected to include these participants to retain power with the relatively smaller sample with available neuroimaging data. Therefore, we used REGNIE which accounts for potential influence of cryptic relatedness (105). This GWAS was performed on SNPs using similar quality control filters as above (INFO > 0.8, minor allele count > 5, minor allele frequency > 1%, missing data

< 10%, and Hardy-Weinberg equilibrium *p* < 1e-6), and covariates included age, genetic sex, batch, and genetic principal components 1-10. We did not observe notable genomic inflation (λ_1000_ = 1.000026). Further details of this GWAS can be found in **Supplemental Information S1.**

All five sets of summary statistics were filtered using MungeSumstats (106). Finally, clumping was performed to identify lead SNPs from independent loci using linkage-disequilibrium scores for European populations (107). Clumping was performed with the following parameters: strict *p* < 5e-8, relaxed *p* < 5e-6, *r^2^* < 0.001, kb window = 10,000.

We performed two sample MR using the inverse-variance weighted method to estimate the effect of sleep phenotypes on DunedinPACNI. We also repeated these analyses using alternative MR estimators (MR-Egger, weighted median, weighted mode, and simple mode) to test the robustness of our findings to potential violations of the instrumental variable assumptions. Horizontal pleiotropic SNPs can violate the exclusion restriction assumption, which requires that the genetic instruments must only affect the outcome through the exposure (39). Therefore, we performed several sensitivity analyses to examine the robustness of our results and assess potential bias due to horizontal pleiotropy. First, we formally tested for heterogeneity among SNP effects by calculating Cochran’s Q statistic. Second, we used the MR-Egger intercept test to assess evidence of directional horizontal pleiotropy. Third, we performed leave-one-out tests by repeating each MR analysis while systematically excluding each instrumental SNP one-at-a-time to test whether any specific SNP disproportionally impacted our results. Fourth, we used MR-PRESSO (108) to identify outlying SNPs that may be pleiotropic and repeated our analyses while excluding these outliers. Fifth, we performed a biologically-grounded leave-many-out analysis where we excluded SNPs that have been found to have genome-wide significant associations with phenotypes that may affect brain structure or aging phenotypes (43). These sensitivity analyses are presented in **Supplemental Information S2-S3.** Finally, we tested for reverse causation (i.e. DunedinPACNI causing sleep phenotypes) using GWAS summary statistics from the UK Biobank insomnia GWAS, the FinnGen and MVP GWAS of sleep disorders, and the FinnGen GWAS of non-organic sleep disorders (**Supplemental Information S2-S3**). It was not possible to test for reverse causation using the 23andMe summary statistics because only the top 10,000 SNPs were available, thus biasing the selection of genetic instruments. All MR analyses were performed using the TwoSampleMR R package.

## DATA AVAILABILITY

Midlife in the United States data are publicly available at https://midus.colectica.org/. Researchers can apply to access all UK Biobank data at https://ams.ukbiobank.ac.uk/ams/. Alzheimer’s Disease Neuroimaging Initiative data are publicly available at https://adni.loni.usc.edu/. Human Connectome Project data is publicly available at https://www.humanconnectome.org/. E-Risk data can be accessed with permission from the E-Risk Study team at https://eriskstudy.com/data-access/. Abbreviated GWAS summary statistics from 23andMe can be accessed at https://cncr.nl/research/summary_statistics/. Full GWAS summary statistics from FinnGen and the Million Veteran Program are publicly available at https://public-mvp-ukbb.finngen.fi/.

## CODE AVAILABILITY

Scripts used in the analyses presented here are available at https://github.com/etw11/Whitman_sleep_2026. The DunedinPACE algorithm is publicly available at https://github.com/danbelsky/DunedinPACE. The DunedinPACNI algorithm is publicly available at https://github.com/etw11/DunedinPACNI.

## ACKNOWLEDGEMENTS

This research received support from the US National Institute on Aging (grants R01AG049789, R01AG032282, R01AG073207) and the UK Medical Research Council (grants MR/X021149/1, MR/X010791). H.L.F. was part supported by the UK Economic and Social Research Council (ESRC) Centre for Society and Mental Health at King’s College London (grants ES/S012567/1, UKRI861). J.M. is funded by the King’s Prize Fellowship and a 2024 NARSAD Young Investigator Grant from the Brain & Behavior Research Foundation (ID 32776). The views expressed are those of the authors and not necessarily those of the ESRC or King’s College London. For the purpose of Open Access, the authors have applied a CC BY public copyright license to any Author Accepted Manuscript version arising from this submission.

This research has been conducted using the UK Biobank Resource under Application Number 493261.

Data collection and sharing for this project was funded by the Alzheimer’s Disease Neuroimaging Initiative (ADNI) (National Institutes of Health Grant U01 AG024904) and DOD ADNI (Department of Defense award number W81XWH-12-2-0012). ADNI is funded by the National Institute on Aging, the National Institute of Biomedical Imaging and Bioengineering, and through generous contributions from the following: AbbVie, Alzheimer’s Association; Alzheimer’s Drug Discovery Foundation; Araclon Biotech; BioClinica, Inc.; Biogen; Bristol-Myers Squibb Company; CereSpir, Inc.; Cogstate; Eisai Inc.; Elan Pharmaceuticals, Inc.; Eli Lilly and Company; EuroImmun; F. Hoffmann-La Roche Ltd and its affiliated company Genentech, Inc.; Fujirebio; GE Healthcare; IXICO Ltd.; Janssen Alzheimer Immunotherapy Research & Development, LLC.; Johnson & Johnson Pharmaceutical Research & Development LLC.; Lumosity; Lundbeck; Merck & Co., Inc.; Meso Scale Diagnostics, LLC.; NeuroRx Research; Neurotrack Technologies; Novartis Pharmaceuticals Corporation; Pfizer Inc.; Piramal Imaging; Servier; Takeda Pharmaceutical Company; and Transition Therapeutics. The Canadian Institutes of Health Research is providing funds to support ADNI clinical sites in Canada. Private sector contributions are facilitated by the Foundation for the National Institutes of Health (www.fnih.org). The grantee organization is the Northern California Institute for Research and Education, and the study is coordinated by the Alzheimer’s Therapeutic Research Institute at the University of Southern California. ADNI data are disseminated by the Laboratory for Neuro Imaging at the University of Southern California.

We are grateful to the E-Risk study mothers and fathers, the twins, and the twins’ teachers and neighbors for their participation. Our thanks to members of the E-Risk team for their dedication, hard work and insights.

