## Supplemental Materials for "Poor sleep is robustly correlated with accelerated aging but the evidence for causation is mixed"

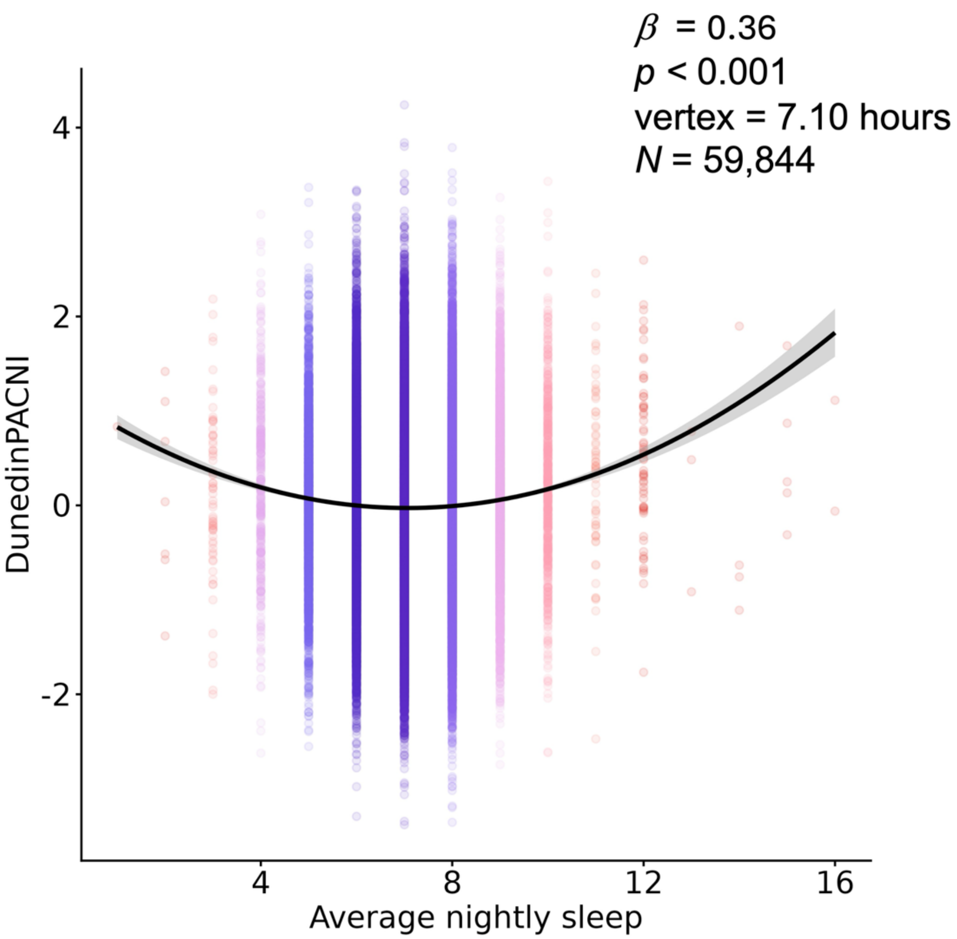

**Supplemental Figure S1. Quadratic association between sleep duration and DunedinPACNI in UK Biobank**. Scatterplot of self-reported average sleep length in hours and DunedinPACNI scores in the UK Biobank. Quadratic regression model controlled for sex and age. Cooler colors represent more normative sleep duration, warmer colors represent less normative sleep duration.

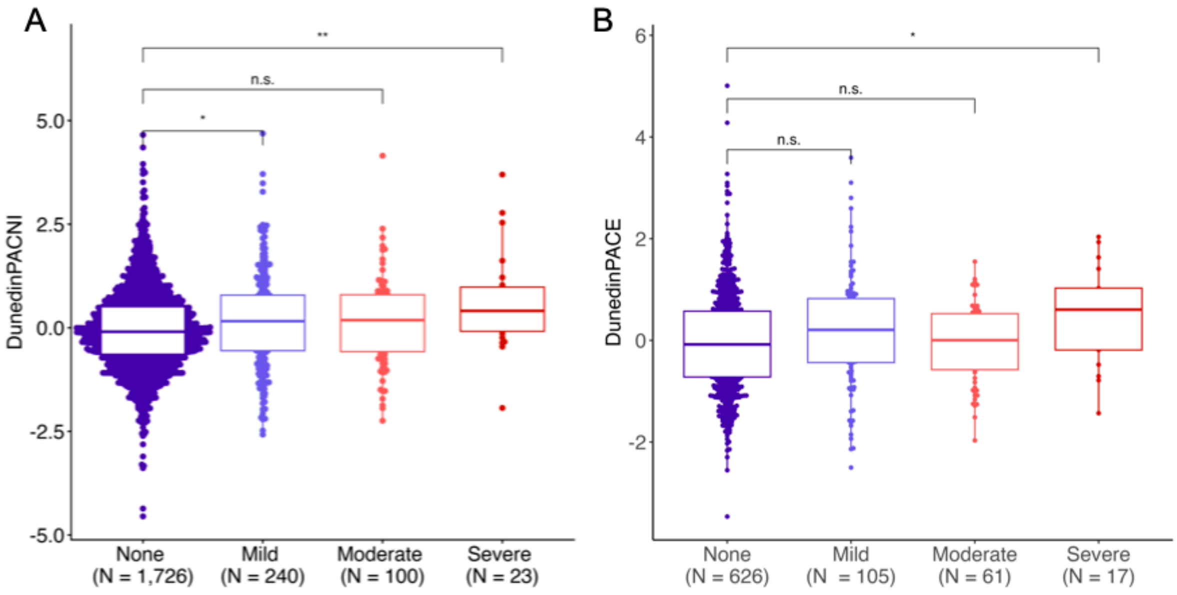

**Supplemental Figure S2. Associations between sleep quality and aging in ADNI participants while controlling for cognitive status. A.** Boxplot of DunedinPACNI scores according to informant-reported sleep impairment among ADNI participants. **B**. Boxplot of DunedinPACE scores according to informant-reported sleep impairment among ADNI participants. Cooler colors represent less sleep impairment; warmer colors represent greater sleep impairment.

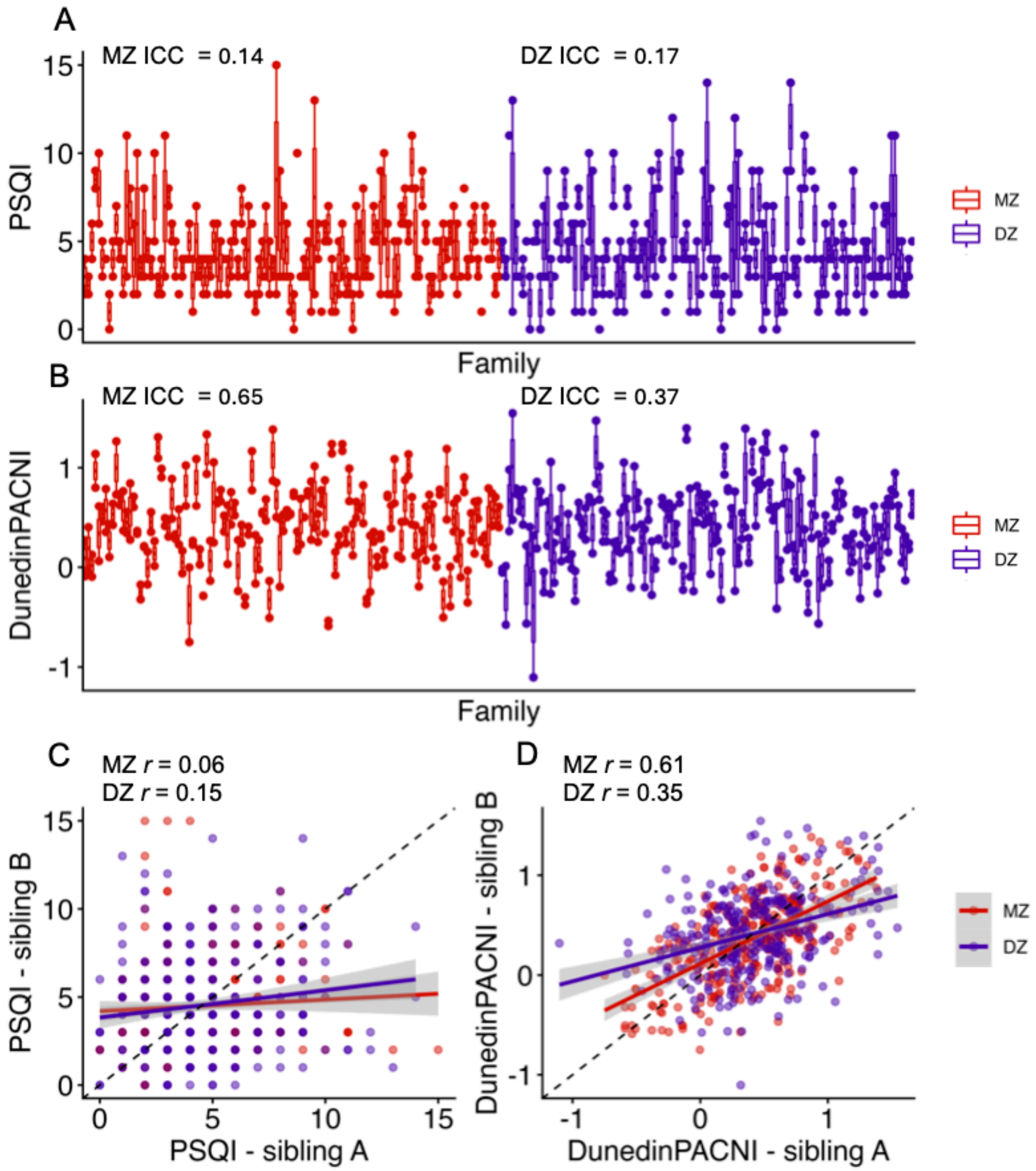

**Supplemental Figure S3. Within-family correlations of PSQI and DunedinPACNI scores in HCP. A.** Boxplot of PSQI global sleep quality scores by family. **B.** Boxplot of DunedinPACNI scores by family. In **A** and **B**, each point represents an individual; individuals are grouped by family along the x-axis. Families are ordered by presence of monozygotic or dizygotic twin pairs in the family. **C.** Scatterplot of correlation in PSQI total sleep quality scores between each pair of twins. **D.** Scatterplot of correlation in DunedinPACNI scores between each pair of twins. In **C** and D, regression lines are presented for correlations within monozygotic and dizygotic twin pairs, respectively. Shaded areas represent 95% confidence intervals. Dotted lines represent y=x. Abbreviations: DZ = dizygotic, HCP = Human Connectome Project, ICC = intraclass correlation, MZ = monozygotic, PSQI = Pittsburgh Sleep Quality Index.

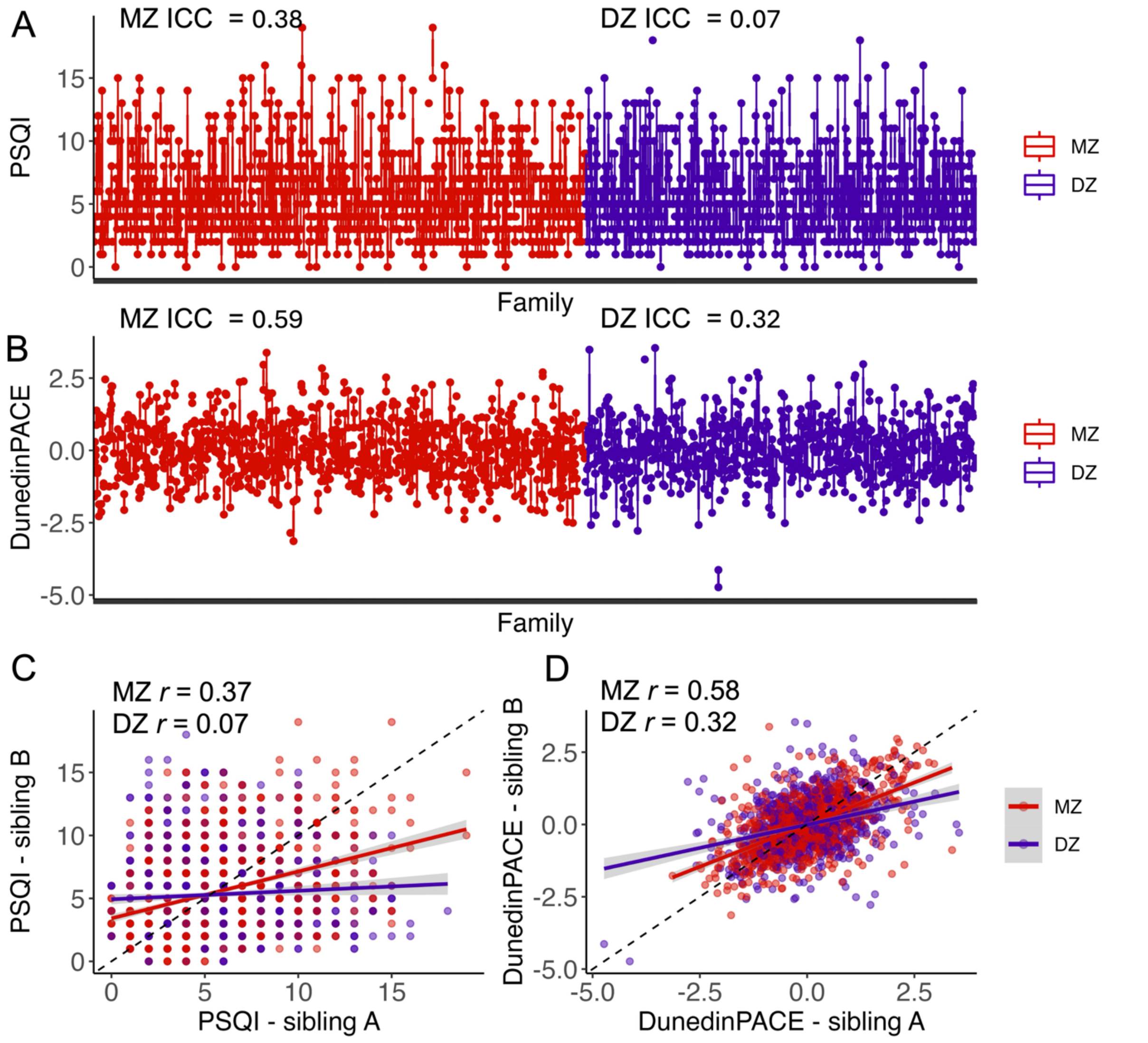

**Supplemental Figure S4. Within-family correlations of PSQI and DunedinPACE scores in E-Risk. A.** Boxplot of PSQI global sleep quality scores by family. **B.** Boxplot of DunedinPACE scores by family. In **A** and **B**, each point represents an individual; individuals are grouped by family along the x-axis. Twins are ordered by zygosity. **C.** Scatterplot of correlation in PSQI total sleep quality scores between each pair of twins. **D.** Scatterplot of correlation in DunedinPACE scores between each pair of twins. In **C** and D, regression lines are presented for correlations within monozygotic and dizygotic twins, respectively. Shaded areas represent 95% confidence intervals. Dotted lines represent y=x. Abbreviations: DZ = dizygotic, ICC = intraclass correlation, MZ = monozygotic, PSQI = Pittsburgh Sleep Quality Index.

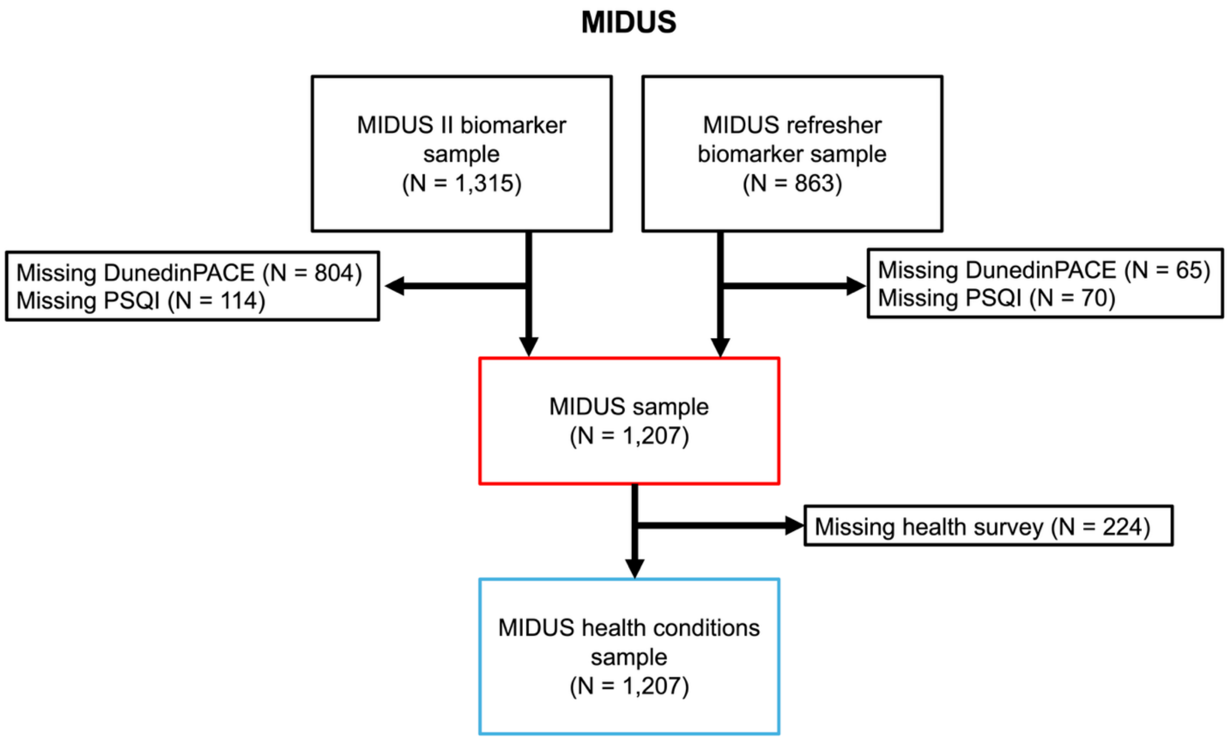

**Supplemental Figure S5. MIDUS Study inclusion flowchart.** Abbreviations: MIDUS = Midlife Development in the United States, PSQI = Pittsburg Sleep Quality Index.

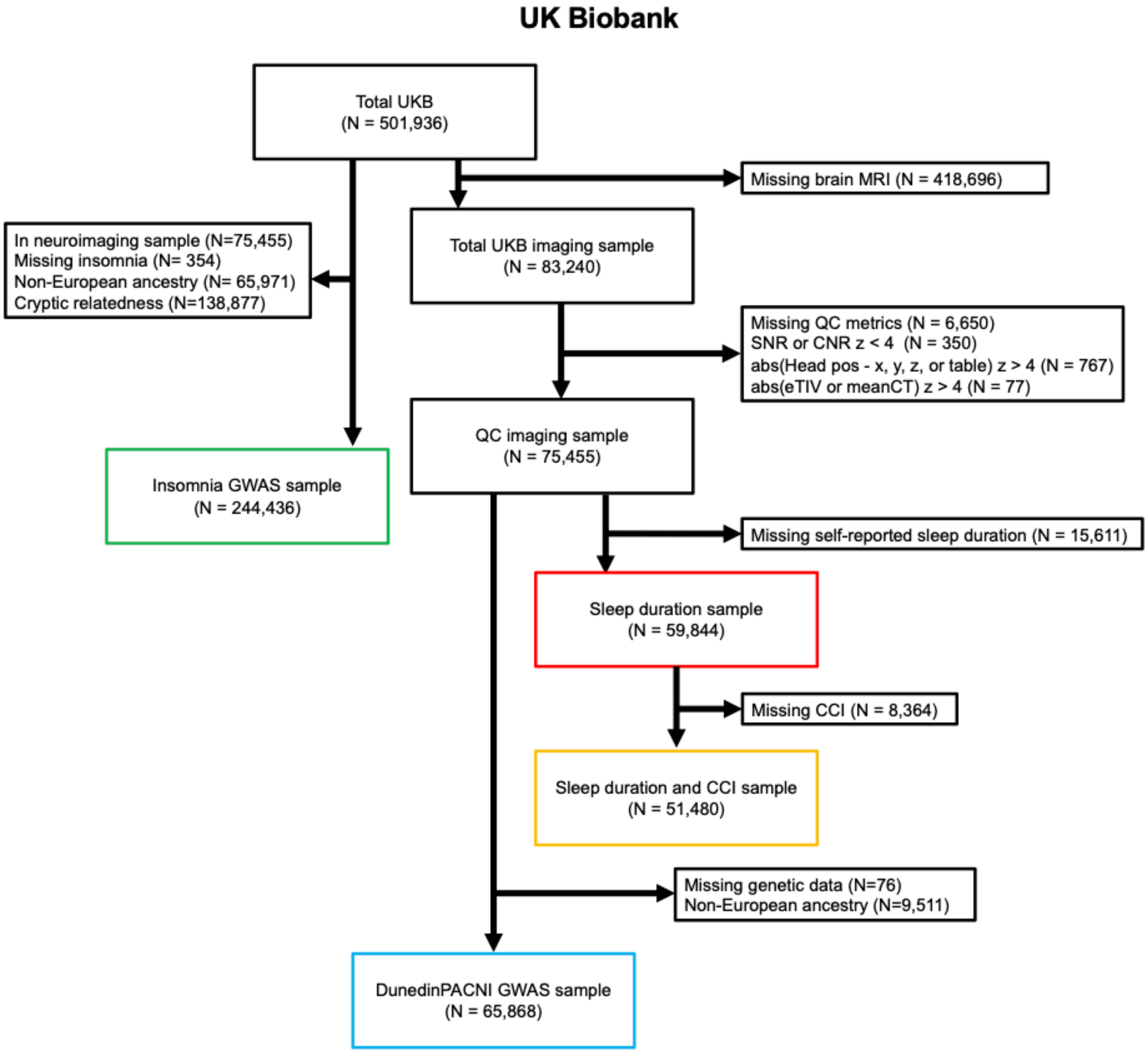

**Supplemental Figure S6. UK Biobank participant inclusion flowchart.** Participants' scans were excluded for low scan signal-to-noise ratio, unusual head positioning, or extremely large intracranial volume or cortical thickness estimates presumed to represent low quality FreeSurfer reconstruction. Abbreviations: CCI = Charlson Comorbidity Index, CNR = contrast-to-noise ratio, eTIV = estimated total intracranial volume, Head pos = head position, meanCT = mean cortical thickness, MRI = magnetic resonance imaging, SNR = signal-to-noise ratio, UKB = UK Biobank.

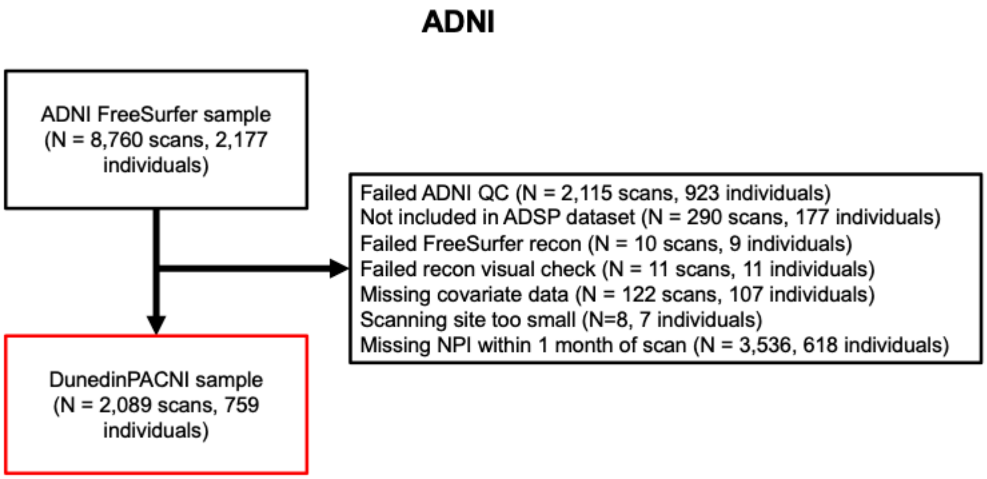

**Supplemental Figure S7. ADNI imaging participant inclusion flowchart.** Participants' scans were deemed to failed ADNI quality control if they were not given a "Pass" according to centralized quality control of FreeSurfer output by ADNI investigators or if their FreeSurfer output failed visual inspection. Abbreviations: ADNI = Alzheimer's Disease Neuroimaging Initiative, ADSP = Alzheimer's Disease Sequencing Project, NPI = Neuropsychiatric Inventory, QC = quality control.

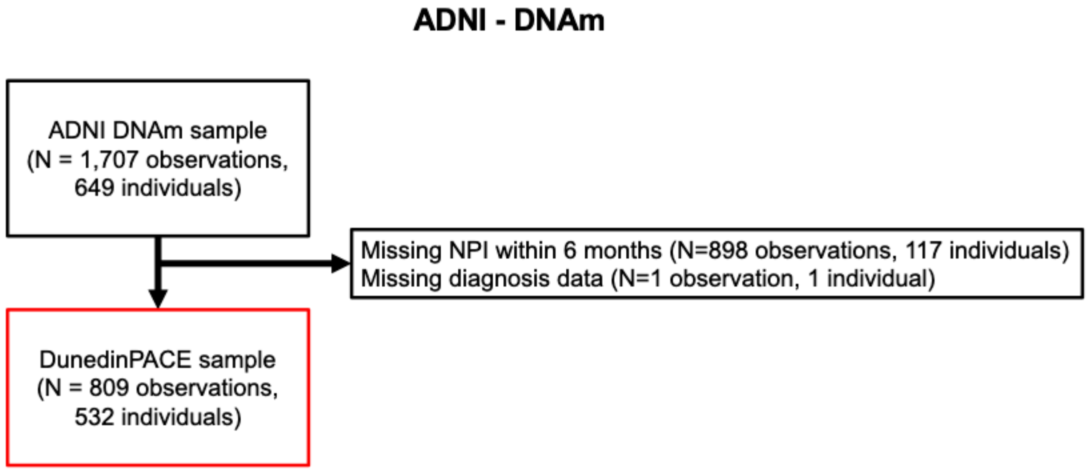

**Supplemental Figure S8. ADNI DNA methylation participant inclusion flowchart.** Abbreviations: ADNI = Alzheimer's Disease Neuroimaging Initiative, DNAm = DNA methylation, NPI = Neuropsychiatric Inventory.

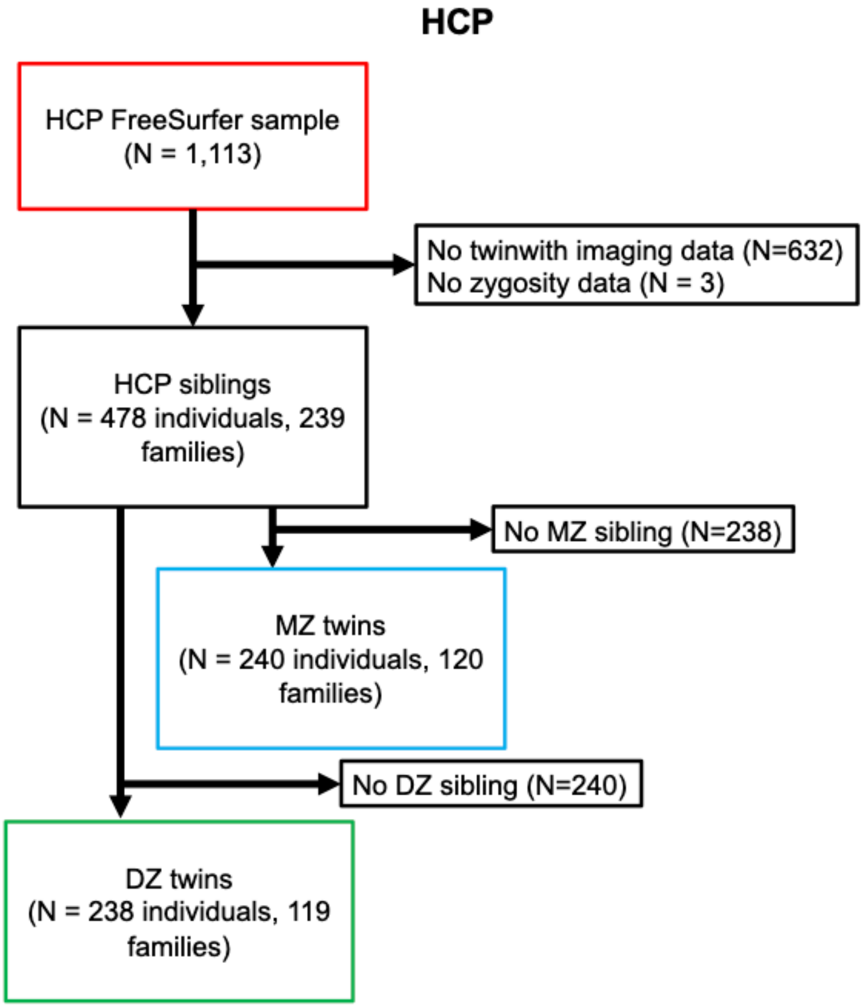

**Supplemental Figure S9. HCP participant inclusion flowchart.** Abbreviations: DZ = dizygotic, HCP = Human Connectome Project, MZ = monozygotic.

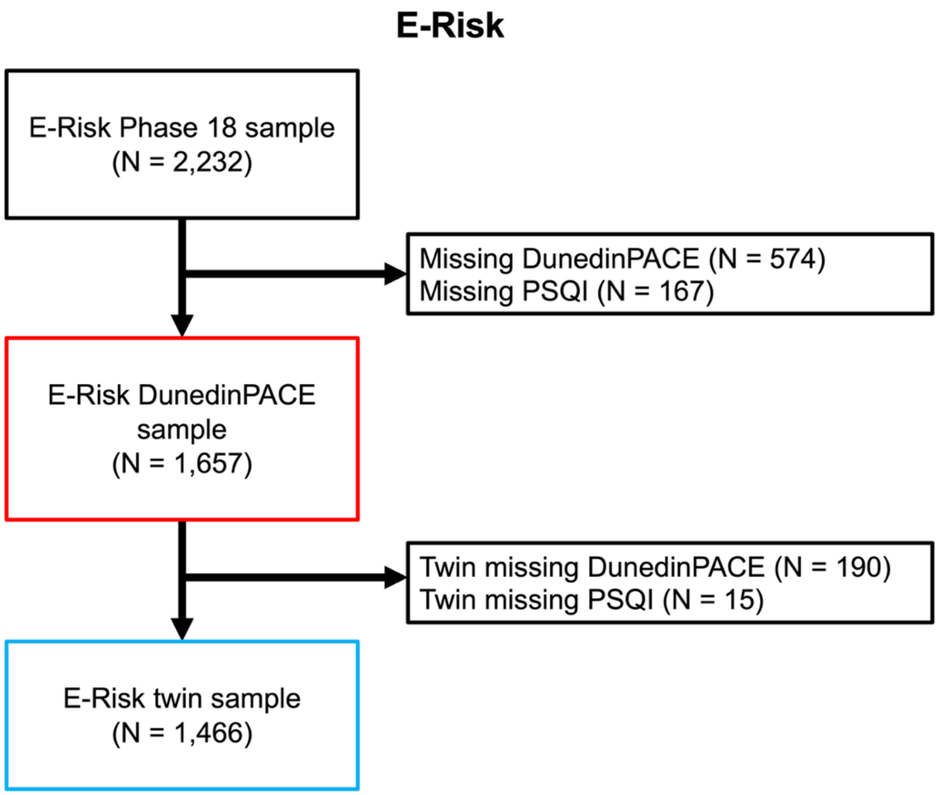

**Supplemental Figure S10. E-Risk participant inclusion flowchart.** Abbreviations: PSQI = Pittsburgh Sleep Quality Index.

**Supplemental Information S1. Detailed genome-wide association analyses**

REGENIE was used to perform genome wide association study (GWAS) analyses (Mbatchou et al. 2021). Briefly, REGENIE performs GWAS in two steps. During step one, a subset of participants' SNP data is used to generate per-individual phenotype predictions using a leave-one-chromosome out approach, serving as a null model. For this step, we used all directly assayed SNPs for all individuals in each GWAS sample. We observed that a small percentage of the DunedinPACNI GWAS sample had cryptic relatedness (4%) according to estimates of genetic kinship (Field ID = 22021). We chose to include these individuals due to their relatively low frequency, the relatively reduced power in the DunedinPACNI GWAS, and because REGENIE effectively accounts for moderate amounts of relatedness and population structure (Mbatchou et al. 2021).

During step two, we tested for associations between imputed SNPs and phenotypes in each GWAS sample, while adjusting for the predictions from step 1. As described in the main text, we excluded SNPs of poor quality (INFO < 0.8, missing > 10%), of low frequency (minor allele frequency < 1%, minor allele count ≤ 5) and SNPs in Hardy-Weinberg disequilibrium ($p$ < 1e-6). Summary statistics were subsequently cleaned using MungeSumstats (Murphy, Schilder, and Skene 2021). Ultimately, we retained associations for 7,101,496 SNPs with insomnia and for 7,101,686 SNPs with DunedinPACNI.

We did not observe notable genomic inflation for either GWAS. To assess inflation, we calculated $\lambda_{1000}$. This is a standard method to assess genomic inflation of polygenic traits which scales the typical $\lambda$ to a sample of 1,000 individuals (Devlin and Roeder 1999; Freedman et al. 2004). This did not indicate evidence of genomic inflation for either GWAS (insomnia: $\lambda_{1000}$ = 1.000029; DunedinPACNI: $\lambda_{1000}$ = 1.000026).

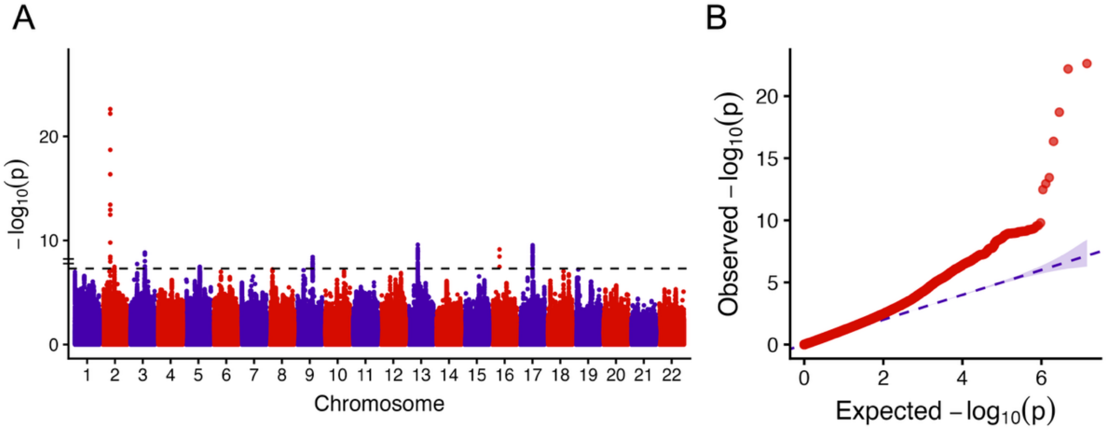

**GWAS of self-reported insomnia in UK Biobank. A.** Manhattan plot of genetic associations with self-reported insomnia. Each point represents a unique SNP, alternating colors indicate different chromosomes. Dashed horizontal line indicates genome-wide significance ($p$ < 5e-8). **B.** QQ plot of genetic associations with self-reported insomnia. Each point represents a unique SNP, dashed line represents equivalence between expected and observed p-values.

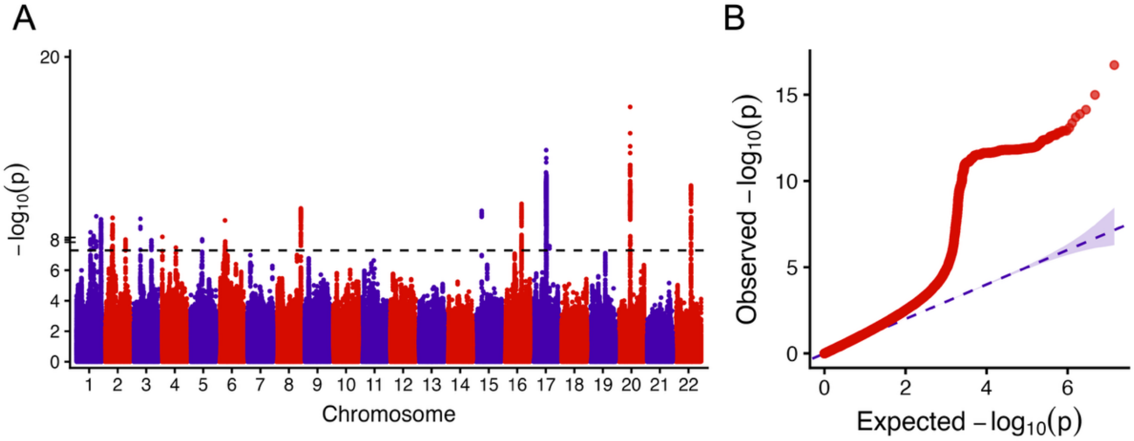

**GWAS of DunedinPACNI in UK Biobank. A.** Manhattan plot of genetic associations with DunedinPACNI. Each point represents a unique SNP, alternating colors indicate different chromosomes. Dashed horizontal line indicates genome-wide significance ($p$ < 5e-8). **B.** QQ plot of genetic associations with self-reported insomnia. Each point represents a unique SNP, dashed line represents equivalence between expected and observed p-values.

**Supplemental Information S2. Mendelian randomization tests of the effect of insomnia on aging.**

We used summary statistics from two GWAS to select genetic instruments for insomnia for Mendelian randomization (MR). The first GWAS was performed in UK Biobank and is described above.

The second GWAS was performed in 23andMe (Watanabe et al. 2022). In this GWAS, insomnia was defined by self-report of seven questions. Participants with positive response to any of the following questions were considered as cases: (i) 'Have you ever been diagnosed with, or treated for, insomnia?', (ii) 'Were you diagnosed with insomnia?', (iii) 'Have you ever been diagnosed by a doctor with any of the following neurological conditions?' (Sleep disturbance), (iv) 'Do you routinely have trouble getting to sleep at night?', (v) 'What sleep disorders have you been diagnosed with? Please select all that apply.' (Insomnia, trouble falling or staying asleep), (vi) 'Have you ever taken these medications?' (Prescription sleep aids) and (vii) 'In the last 2 years, have you taken any of these medications?' (Prescription sleep aids). Participants who did not provide either positive or uncertain answer ('I don't know' or 'I am not sure') in any of the seven questions nor the followings are considered as controls: (i) 'Have you ever been diagnosed with, or treated for, any of the following conditions?' (Insomnia; Narcolepsy; Sleep apnea; Restless leg syndrome), (ii) 'In the past 12 months, have you been newly diagnosed with any of the following conditions by a medical professional?' (Insomnia; Sleep apnea; Migraines), (iii) 'Have you ever been diagnosed with or treated for any of the following conditions?' (Posttraumatic stress disorder; Autism; Asperger's; Sleep disorder), (iv) 'Have you ever been diagnosed with or treated for a sleep disorder?', and (v) 'Have you ever been diagnosed with or treated for any of the following conditions?' (A sleep disorder), (Watanabe et al. 2022). The top 10,000 SNPs from this GWAS are publicly available. These GWAS summary statistics were also processed using MungeSumstats as described above.

After quality control of GWAS summary statistics, we performed a clumping analysis to identify lead SNPs that would be suitable genetic instruments. This clumping analysis identified lead SNPs with $r^2 < 0.001$ and outside of a 10,000kb window. We chose relatively strict clumping parameters to ensure independence of genetic instruments in Mendelian randomization tests. Lead SNPs were first considered among genome-wide significant SNPs ($p$ < 5e-8). SNPs identified as instrumental variables for insomnia are presented in **Individual SNP Level Tables**. Mendelian randomization was performed using the *TwoSampleMR* package in R.

1. *Effect of insomnia on DunedinPACNI with instrumental SNPs from the UK Biobank insomnia GWAS*

We first performed Mendelian randomization to test the effects of insomnia on DunedinPACNI using genetic instruments identified from the GWAS of self-reported insomnia in UK Biobank. We did not find evidence for a causal effect of insomnia on DunedinPACNI from inverse-variance weighted meta-analytic testing. Additional tests were also null.

**MR tests of the effect of insomnia on DunedinPACNI using instrumental SNPs from the UK Biobank GWAS.**

| Exposure | Exposure instrument | Outcome | Outcome instrument | Method | *N* SNP | *b* | *p* |
|---|---|---|---|---|---|---|---|
| Insomnia | UKB non-imaging GWAS | DunedinPACNI | UKB imaging GWAS | MR Egger | 9 | -0.39 | 0.33 |
| | | | | Weighted median | 9 | -0.14 | 0.19 |
| | | | | Inverse variance weighted | 9 | -0.09 | 0.50 |
| | | | | Simple mode | 9 | -0.10 | 0.53 |
| | | | | Weighted mode | 9 | -0.14 | 0.43 |

We observed heterogeneity between these instrumental SNPs (Q = 21.34, $p = 0.003$), though we did not observe evidence of directional horizontal pleiotropy (MR-Egger intercept = 0.005, $p = 0.42$). We visualized effects of each SNP using a forest plot. To further assess the robustness of our findings, we performed a leave-one-out analysis for each instrumental SNP. We observed generally consistent null effects, though we did find that the effect of *rs4688760* appeared to be pulling the overall effect toward the null.

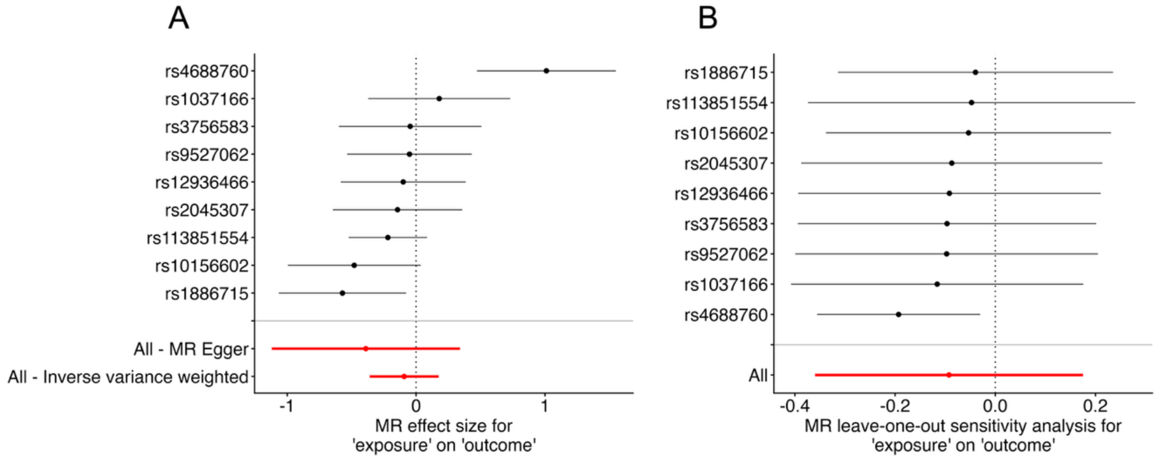

**Sensitivity tests of UK Biobank insomnia GWAS MR analyses. A.** Forest plot of SNP effects of insomnia on DunedinPACNI. **B.** Forest plot of leave-one-out effects of insomnia on DunedinPACNI. Each row represents the row when that SNP is left out.

We formally tested for and corrected for horizontal pleiotropy effects in this analysis using MR-PRESSO (Verbanck et al. 2018). The MR-PRESSO global test identified evidence of horizontal pleiotropy ($p = 0.004$) for *rs4688760*. The outlier-corrected inverse variance weighted estimate was substantially larger than the original estimate (outlier-corrected: $\beta$=-0.19, $p$=0.04; original: $\beta$=-0.09, $p$=0.50) and statistically significant, however, we note that this effect is in the opposite direction (i.e., that insomnia causes slower aging).

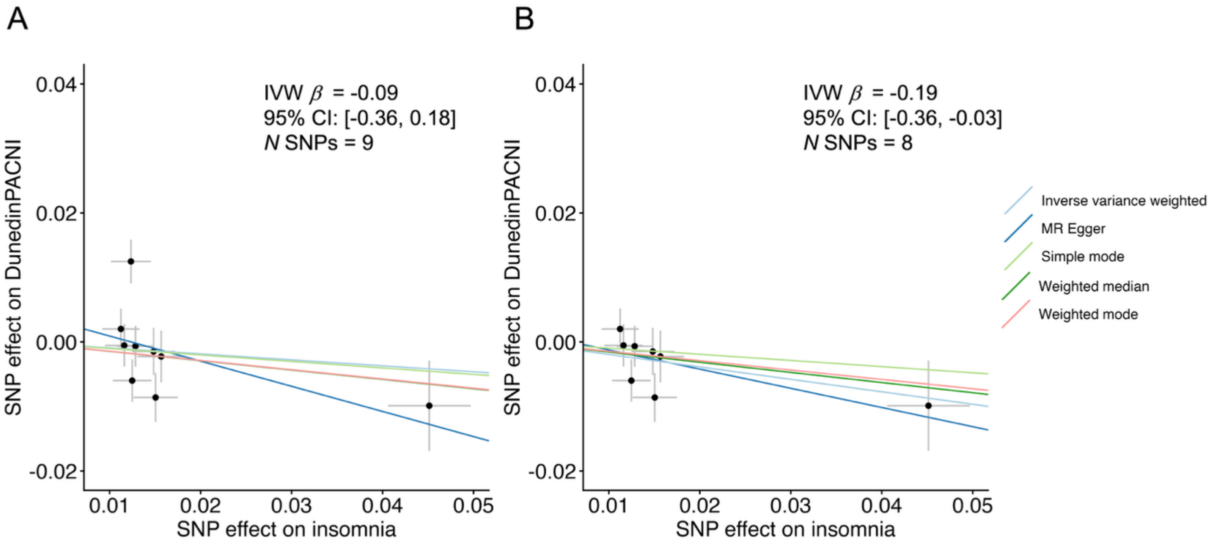

**MR-PRESSO outlier-correction of the effect of insomnia on DunedinPACNI. A.** Original model using all 9 instrumental SNPs from the UK Biobank insomnia GWAS. **B.** Model after using MR-PRESSO to remove outlying SNPs.

Finally, we performed a biologically grounded sensitivity test for horizontal pleiotropy. Sleep is a complex behavior that is influenced by many pleiotropic alleles, thus necessitating careful examination of potential pleiotropic bias in MR analyses (Evans et al. 2025). To do this, we searched the NHGRI-EBI GWAS Catalog to find other genome-wide significant associations for instrumental SNPs using the *gwasrapidd* R package (Magno and Maia 2020). We then performed a "leave-many-out" analysis by excluding SNPs that had associations with any neuroimaging phenotypes or with phenotypes used in the generation of the original Pace of Aging (Elliott et al. 2021), as these phenotypes may affect DunedinPACNI scores.

We excluded four SNPs (*rs113851554, rs4688760, rs10156602,* and *rs12936466*) associated with potentially confounding phenotypes (see full list in **Individual SNP Level Tables**). We again observed a null effect of insomnia on DunedinPACNI (leave-many-out: $\beta$=-0.14, $p$=0.25, original: $\beta$=-0.09, $p$=0.50).

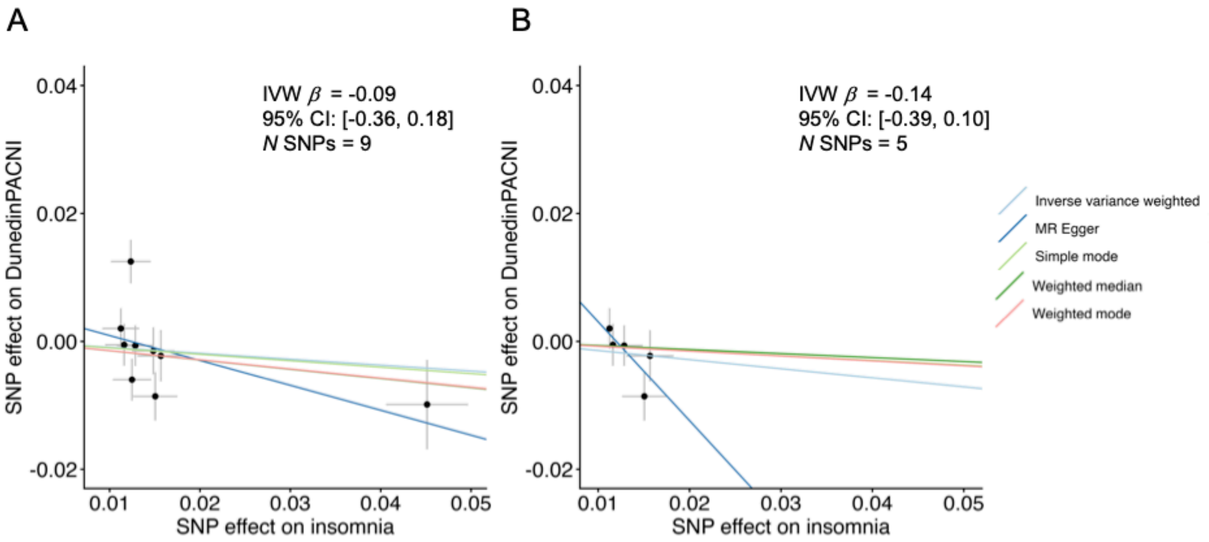

**Testing the effect of SNPs associated with confounding phenotypes on the causal influence of insomnia on DunedinPACNI. A.** Original model using all 9 instrumental SNPs from the 23andMe insomnia GWAS. **B.** Model after removing potentially confounded SNPs.

Taken together, this analysis provided no evidence for a causal association between insomnia, as measured via self-report, and accelerated aging. Sensitivity analyses identified model instability and potential bias due to horizontal pleiotropy and a low number of instrumental SNPs. Therefore, we attempted to replicate this finding using instrumental SNPs from the GWAS of insomnia conducted in 23andMe.

2. *Effect of insomnia on DunedinPACNI with instrumental SNPs from the 23andMe insomnia GWAS*

Using summary statistics from the 23andMe insomnia GWAS, we again did not find strong evidence for a causal effect of insomnia on DunedinPACNI from inverse-variance weighted meta-analytic testing. Additional tests were mostly null with the notable exception of the weighted mean.

**MR tests of the effect of insomnia on DunedinPACNI using instrumental SNPs from the 23andMe GWAS.**

| Exposure | Exposure instrument | Outcome | Outcome instrument | Method | *N* SNP | *b* | *p* |
|---|---|---|---|---|---|---|---|
| Insomnia | 23andMe GWAS | DunedinPACNI | UKB imaging GWAS | MR Egger | 105 | 0.03 | 0.78 |
| | | | | Weighted median | 105 | 0.06 | 0.01 |
| | | | | Inverse variance weighted | 105 | 0.03 | 0.16 |
| | | | | Simple mode | 105 | 0.13 | 0.12 |
| | | | | Weighted mode | 105 | 0.12 | 0.15 |

We observed heterogeneity between these instrumental SNPs (Q = 296.94, $p < 0.001$), though we did not observe evidence of directional horizontal pleiotropy (MR-Egger intercept = 0.0002, $p = 0.94$). We again visualized effects of each SNP using a forest plot. To further assess the robustness of our findings, we first performed a leave-one-out analysis for each instrumental SNP. We observed consistent effects when each individual SNP was left out, suggesting that our findings are not driven by the effect of a single instrumental SNP.

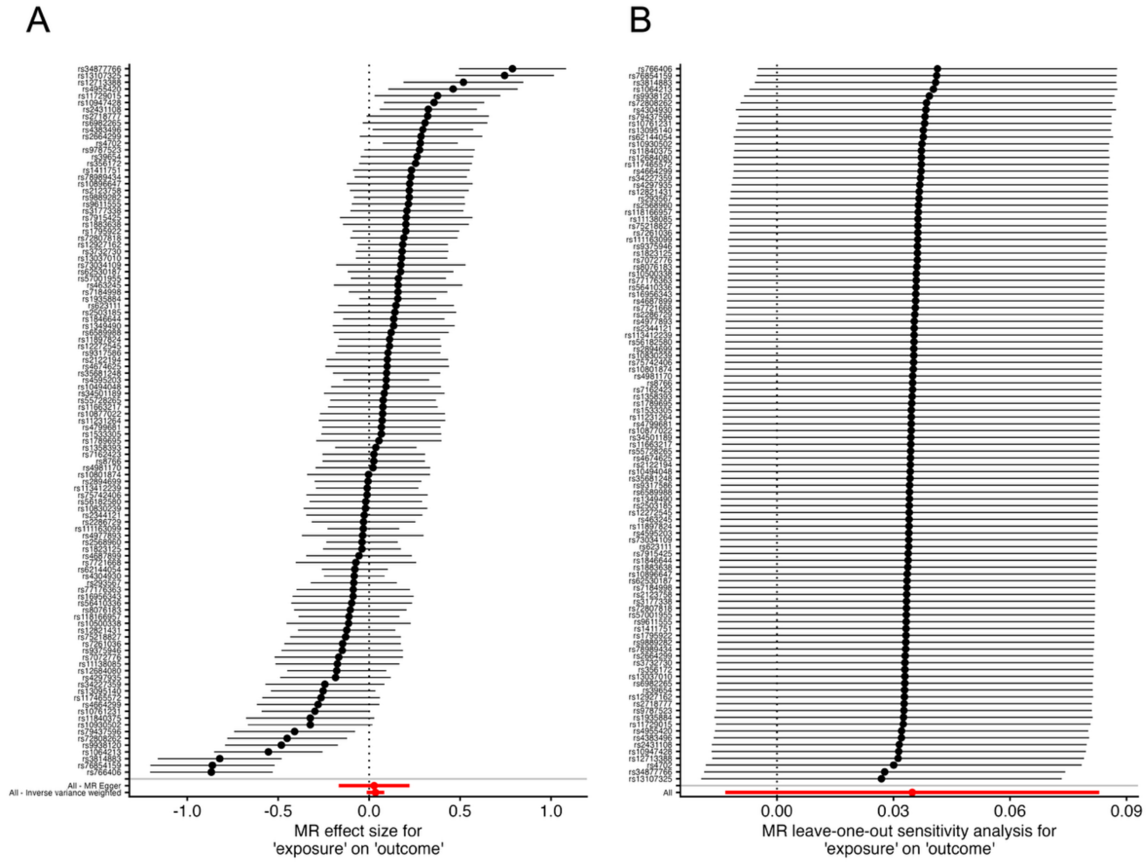

**Sensitivity tests of 23andMe insomnia GWAS MR analyses. A.** Forest plot of SNP effects of insomnia on DunedinPACNI. **B.** Forest plot of leave-one-out effects of insomnia on DunedinPACNI. Each row represents the row when that SNP is left out.

We again formally tested for and corrected for horizontal pleiotropy effects in this analysis using MR-PRESSO (Verbanck et al. 2018). The MR-PRESSO global test identified evidence of horizontal pleiotropy ($p < 0.0041$) driven by *rs3814883, rs34877766, rs76854159, rs766406, rs1064213,* and *rs13107325*. The outlier-corrected inverse variance weighted causal estimate was slightly stronger than the original estimate and statistically significant (Outlier-corrected: $\beta$=0.04, $p$=0.02; original: $\beta$=0.03, $p$=0.16), suggesting that a subset of horizontally pleiotropic SNPs may have drawn the effect towards the null. Of note, however, is that some MR estimators showed effects in the opposite direction after removing outlying SNPs, somewhat reducing the confidence in the robustness of this result.

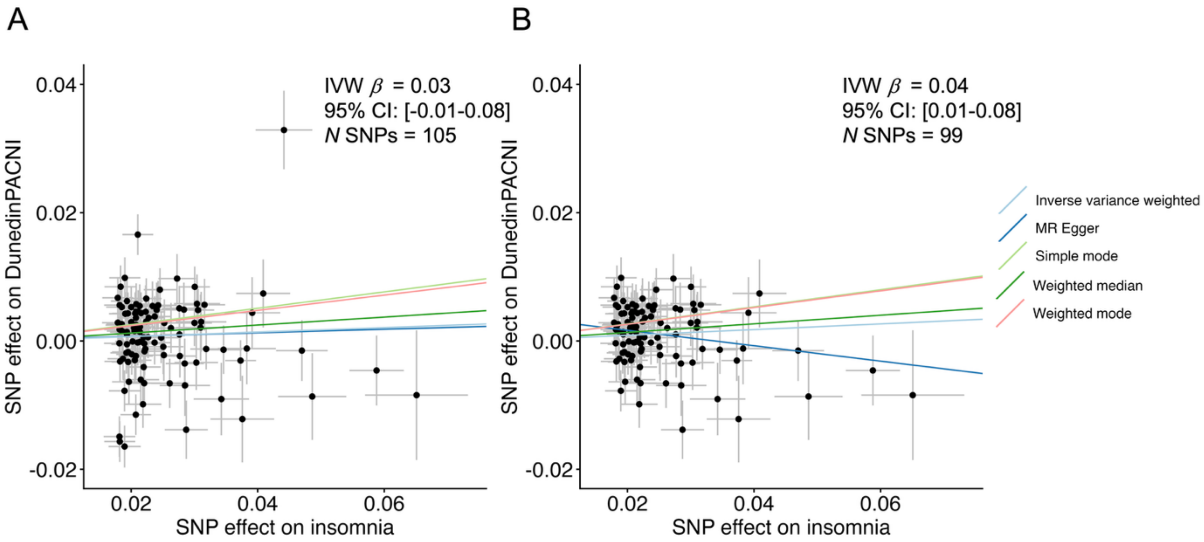

**MR-PRESSO outlier-correction of the effect of insomnia on DunedinPACNI. A.** Original model using all 105 instrumental SNPs from the 23andMe insomnia GWAS. **B.** Model after using MR-PRESSO to remove outlying SNPs.

Finally, we again performed a biologically-grounded "leave-many-out" analysis by excluding SNPs that had associations with any neuroimaging phenotypes or with phenotypes used in the generation of the original Pace of Aging (Elliott et al. 2021), as these phenotypes may affect DunedinPACNI scores.

We excluded 16 SNPs associated with potentially confounding phenotypes (see full list in **Individual SNP Level Tables**). We observed a slightly stronger and statistically significant effect of insomnia on DunedinPACNI after these exclusions (leave-many-out: $\beta$=0.05, $p$=0.04, original: $\beta$=0.03, $p$=0.16).

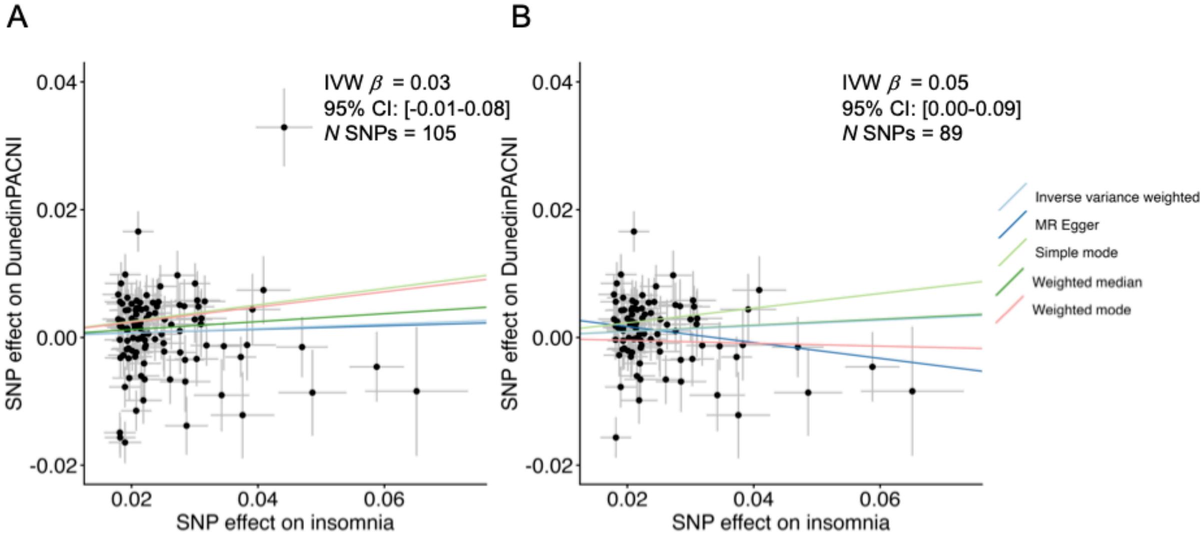

**Testing the effect of SNPs associated with confounding phenotypes on the causal influence of insomnia on DunedinPACNI. A.** Original model using all 105 instrumental SNPs from the 23andMe insomnia GWAS. **B.** Model after removing potentially confounded SNPs.

Collectively, these results present limited evidence for a causal influence of insomnia on faster aging. Although some sensitivity tests were statistically significant while using instruments from the 23andMe GWAS, these results were not consistent with analyses using instruments from the UK Biobank GWAS. Further, some sensitivity tests using SNPs from the UK Biobank GWAS showed effects in the opposite direction, reducing the strength of evidence for a casual effect.

3. *Effect of DunedinPACNI on insomnia with instrumental SNPs from the UK Biobank DunedinPACNI GWAS*

Finally, we tested for casual influence of aging on insomnia. We identified 20 instrumental SNPs for DunedinPACNI from the GWAS described above, shown in **Individual SNP Level Tables**. We first tested the influence of DunedinPACNI on insomnia using the summary statistics from the insomnia GWAS performed in UK Biobank (described above). This analysis did not show evidence of a causal effect (inverse-variance weighted $\beta$=0.04, $p$ = 0.21, 95% CI = [-0.02-0.11], for other methods see table below.

**MR tests of the effect of DunedinPACNI on insomnia using instrumental SNPs from the UK Biobank GWAS.**

| Exposure | Exposure instrument | Outcome | Outcome instrument | Method | *N* SNP | *b* | *p* |
|---|---|---|---|---|---|---|---|
| DunedinPACNI | UKB imaging GWAS | Insomnia | UKB non-imaging GWAS | MR Egger | 20 | -0.08 | 0.61 |
| | | | | Weighted median | 20 | 0.05 | 0.14 |
| | | | | Inverse variance weighted | 20 | 0.04 | 0.21 |
| | | | | Simple mode | 20 | 0.12 | 0.45 |
| | | | | Weighted mode | 20 | 0.11 | 0.50 |

We observed heterogeneity between these instrumental SNPs (Q = 40.52, $p$ = 0.002), though we did not observe evidence of directional pleiotropy (MR-Egger intercept = 0.004, $p$ = 0.43). Leave-one-out analysis did not reveal evidence that this result was driven by any one instrumental SNP.

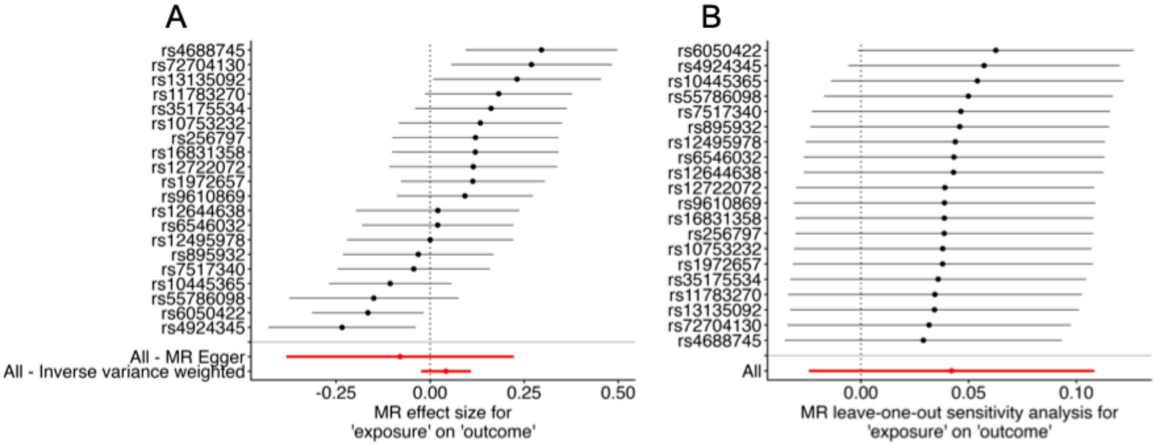

**Sensitivity tests of DunedinPACNI GWAS MR analyses using UK Biobank insomnia GWAS. A.** Forest plot of SNP effects of DunedinPACNI on insomnia. **B.** Forest plot of leave-one-out effects of DunedinPACNI on insomnia. Each row represents the row when that SNP is left out.

We again formally tested for horizontal pleiotropy effects using MR-PRESSO. This analysis identified evidence of horizontal pleiotropy ($p$ = 0.001) that was driven by *rs4688760*. The outlier-corrected inverse variance weighted causal estimate was slightly stronger than the original estimate (Outlier-corrected: $\beta$=0.06, $p$=0.06; original: $\beta$=0.04, $p$=0.21). Of note, however, is that some MR estimators showed effects in opposite directions after removing the outlying SNP, reducing the confidence in the robustness of this result.

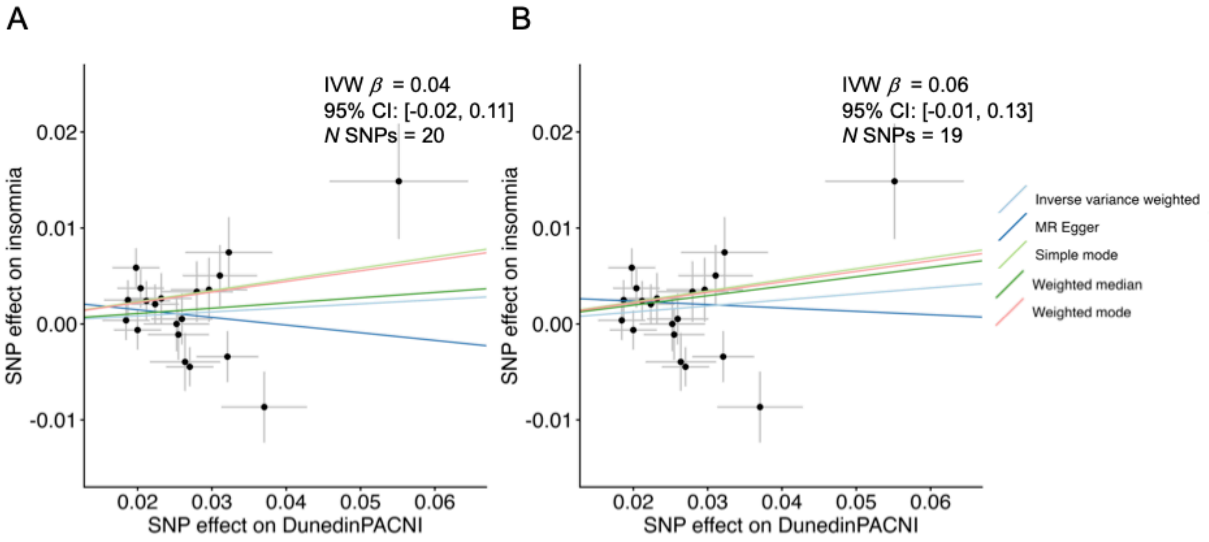

**MR-PRESSO outlier-correction of the effect of DunedinPACNI on insomnia. A.** Original model using all 20 instrumental SNPs from the DunedinPACNI GWAS. **B.** Model after using MR-PRESSO to remove the outlying SNP.

We were unable to test the effect of DunedinPACNI on insomnia using the GWAS from 23andMe because only the top 10,000 SNPs from that GWAS are available. This would bias MR estimates because instrument-outcome associations could only be obtained for SNPs highly associated with insomnia.

Collectively, these analyses provide limited evidence for a causal effect of DunedinPACNI on self-reported insomnia.

**Supplemental Information S3. Mendelian randomization tests of the effect of clinical sleep disorders on aging.**

We further used summary statistics from two GWAS to select genetic instruments for sleep disorders alongside the above GWAS of DunedinPACNI to perform MR. The first sleep disorder GWAS was a meta-analysis of associations from FinnGen and the Million Veterans Program. Cases for this GWAS had a diagnosis of an organic or a non-organic sleep disorder in primary care, hospital discharge, or death reports. Organic sleep disorders are considered neurological or physiological in origin, whereas non-organic sleep disorders are considered psychological in origin and are not caused by an identifiable physical disorder.

The specific diagnoses included as organic sleep disorders were from ICD-10 category G47 (insomnia, hypersomnia, disorders of the sleep-wake schedule, sleep apnea, narcolepsy and cataplexy, other sleep disorders, or sleep disorder, unspecified. The specific diagnoses included as non-organic sleep disorders were from ICD-10 category F51 (insomnia, hypersomnia, disorder of the sleep-wake schedule, sleepwalking, sleep terrors, nightmares, other nonorganic sleep disorders, or nonorganic sleep disorder, unspecified). Controls did not have any of the above diagnoses, nor did they have a diagnosis of any behavioral syndromes associated with physiological disturbances [e.g. eating disorders (ICD-10 category F50), sexual dysfunction (ICD-10 category F52), postpartum depression (ICD-10 category F53), abuse of non-dependence-producing substances (ICD-10 category F55), or psychological and behavioral factors associated with disorders or diseases classified elsewhere (ICD-10 category F54). Further details of this GWAS can be found at (https://public-mvp-ukbb.finngen.fi/).

Variants with imputation quality $r^2 < 0.6$ were excluded; in MVP, additional filters were applied for allele frequency differences > 0.2 compared to gnomAD, a large-scale aggregation of human exome and genome sequencing data. Heterogeneity was assessed with Cochran's Q test. We derived cohort-specific sample sizes (FinnGen, MVP EUR, MVP AFR and MVP HIS) from case/control counts (https://mvp-ukbb.finngen.fi). Weighted allele counts were summed to derive a combined allele frequency and only variants with an allele frequency $\in [0.01, 0.99]$ and standard alleles (A/C/G/T) were retained. These GWAS summary statistics were also processed using MungeSumstats as described above.

The second sleep disorder GWAS was performed in FinnGen only. This GWAS only included non-organic sleep disorders to more closely isolate associations with poor sleep in the absence of physical illness. Cases for this GWAS had a diagnosis of a non-organic sleep disorder from ICD-10 category F51 (insomnia, hypersomnia, disorder of the sleep-wake schedule, sleepwalking, sleep terrors, nightmares, other nonorganic sleep disorders, or nonorganic sleep disorder, unspecified). Controls did not have any of these diagnoses nor did they have a diagnosis of any psychiatric or neurological disorder, as these may affect sleep. These disorders included dementia (ICD-10 categories F00-F03), delirium superimposed on dementia (ICD-10 code F05.1), other degenerative diseases of the nervous system (ICD-10 category G30), any other mental or behavioral disorders (ICD-10 categories F1-F9), or intentional self-harm (ICD-10 categories X60-X84). Further details of this GWAS can be found at (https://public-mvp-ukbb.finngen.fi/). These GWAS summary statistics were also processed using MungeSumstats as described above.

After quality control of GWAS summary statistics, we performed a clumping analysis to identify lead SNPs that would be suitable genetic instruments for MR. This clumping analysis identified lead SNPs with $r^2 < 0.001$ and outside of a 10,000kb window. We chose relatively strict clumping parameters to ensure independence of genetic instruments in Mendelian randomization tests. Lead SNPs were first considered among genome-wide significant SNPs ($p$ < 5e-8), however, in the FinnGen GWAS of nonorganic sleep disorders, no SNPs were significant at that threshold. Therefore, we relaxed this threshold to $p$ < 5e-6 to identify genetic instruments, as is standard practice in MR analysis (Hu et al. 2024). Only three instrumental SNPs (5.9% of total) from the FinnGen and MVP GWAS meta-analysis showed evidence of meta-analytic heterogeneity ($p$ < 0.01). SNPs identified as instrumental variables for sleep disorders are presented in **Individual SNP Level Tables**. Mendelian randomization was performed using the *TwoSampleMR* package in R.

1. *Effect of sleep disorders on DunedinPACNI with instrumental SNPs from the FinnGen and MVP sleep disorder GWAS*

We first performed Mendelian randomization to test the effects of sleep disorders on DunedinPACNI using genetic instruments identified from the GWAS meta-analysis of sleep disorders in FinnGen and MVP. We found evidence for a causal effect of sleep disorders on DunedinPACNI from inverse-variance weighted meta-analytic testing ($\beta$= 0.07, $p$=0.02, 95% CI: 0.01-0.12). Additional tests were in the same direction and of a similar magnitude.

**MR tests of the effect of sleep disorders on DunedinPACNI using instrumental SNPs from the FinnGenn and MVP GWAS.**

| Exposure | Exposure instrument | Outcome | Outcome instrument | Method | *N* SNP | *b* | *p* |
|---|---|---|---|---|---|---|---|
| Sleep disorders | FinnGen + MVP GWAS | DunedinPACNI | UKB imaging GWAS | MR Egger | 52 | 0.07 | 0.41 |
| | | | | Weighted median | 52 | 0.07 | 0.02 |
| | | | | Inverse variance weighted | 52 | 0.07 | 0.02 |
| | | | | Simple mode | 52 | 0.11 | 0.25 |
| | | | | Weighted mode | 52 | 0.10 | 0.30 |

We observed heterogeneity between these instrumental SNPs (Q = 119.55, $p$ < 0.001), though we did not observe evidence of directional horizontal pleiotropy (MR-Egger intercept = -0.0001, $p$ = 0.96). We visualized effects of each SNP using a forest plot. To further assess the robustness of our findings, we performed a leave-one-out analysis for each instrumental SNP. We observed consistent effects across leave-one-out tests, suggesting that this result was not driven by any specific SNP.

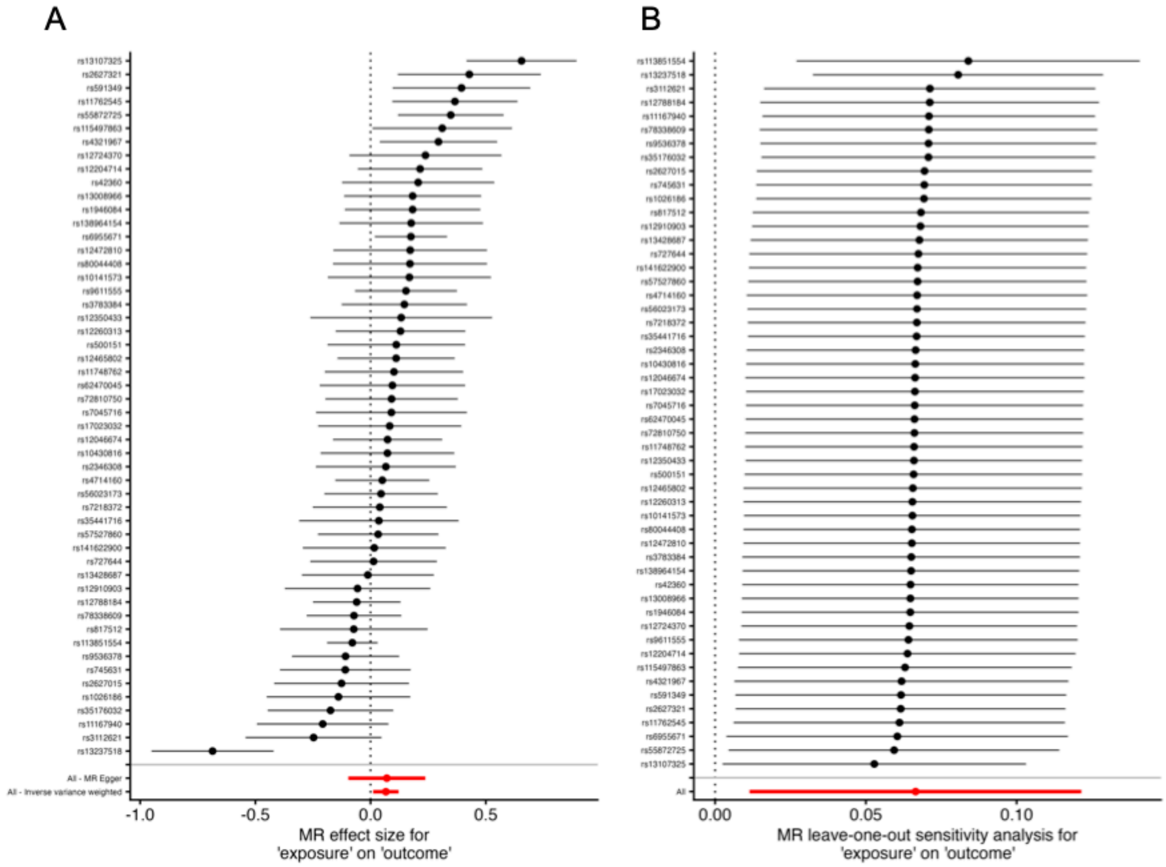

**Sensitivity tests of FinnGen and MVP sleep disorder GWAS meta-analysis MR analyses. A.** Forest plot of SNP effects of sleep disorders on DunedinPACNI. **B.** Forest plot of leave-one-out effects of sleep disorders on DunedinPACNI. Each row represents the row when that SNP is left out.

We formally tested for and corrected for horizontal pleiotropy effects in this analysis using MR-PRESSO (Verbanck et al. 2018). The MR-PRESSO global test identified evidence of horizontal pleiotropy ($p < 0.001$) driven by *rs13107325, rs13237518.* The outlier-corrected inverse variance weighted causal estimate was highly similar than the original estimate (Outlier-corrected: $\beta$=0.07, $p$=0.002; original: $\beta$=0.07, $p$=0.02), suggesting that horizontally pleiotropic SNPs may have drawn the effect towards the null.

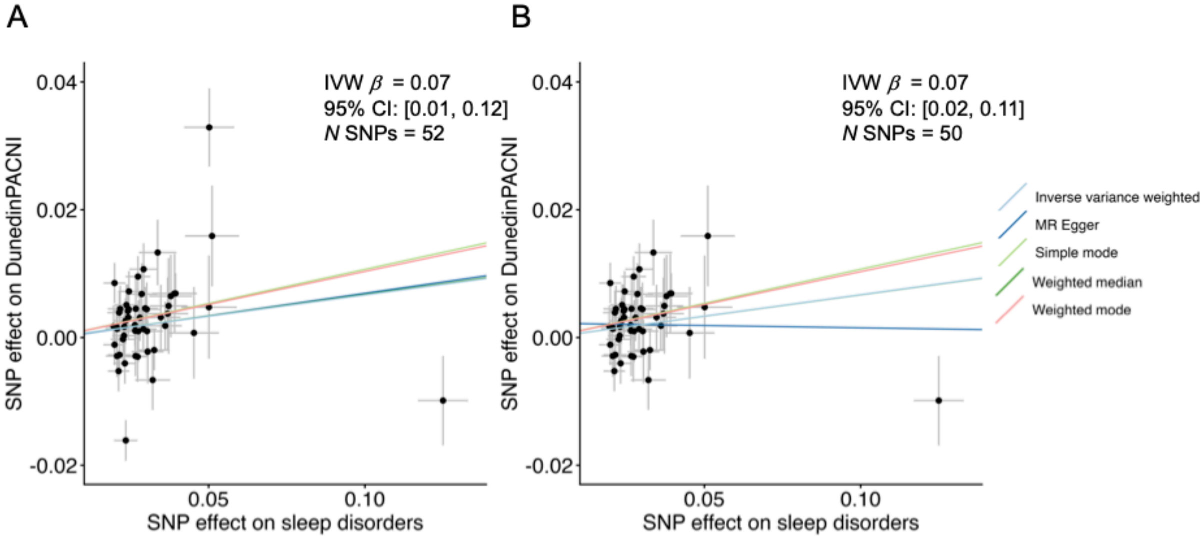

**MR-PRESSO outlier-correction of the effect of sleep disorders on DunedinPACNI. A.** Original model using all 52 instrumental SNPs from the FinnGen and MVP sleep disorder GWAS. **B.** Model after using MR-PRESSO to remove outlying SNPs.

Finally, we performed a biologically grounded sensitivity test for horizontal pleiotropy. Sleep is a complex behavior that is influenced by many pleiotropic alleles, thus necessitating careful examination of potential pleiotropic bias in MR analyses (Evans et al. 2025). To do this, we searched the NHGRI-EBI GWAS Catalog to find other genome-wide significant associations for instrumental SNPs using the *gwasrapidd* R package (Magno and Maia 2020). We then performed a "leave-many-out" analysis by excluding SNPs that had associations with any neuroimaging phenotypes or with phenotypes used in the generation of the original Pace of Aging (Elliott et al. 2021), as these phenotypes may affect DunedinPACNI scores.

We excluded six SNPs (*rs13107325, rs6955671, rs4321967, rs55872725, rs141622900*, and *rs45551238*) associated with potentially confounding phenotypes (see full list in **Individual SNP Level Tables**). We observed a marginally weaker effect of sleep disorders on DunedinPACNI after these exclusions (leave-many-out: $\beta$=0.03, $p$=0.23, original: $\beta$=0.07, $p$=0.02).

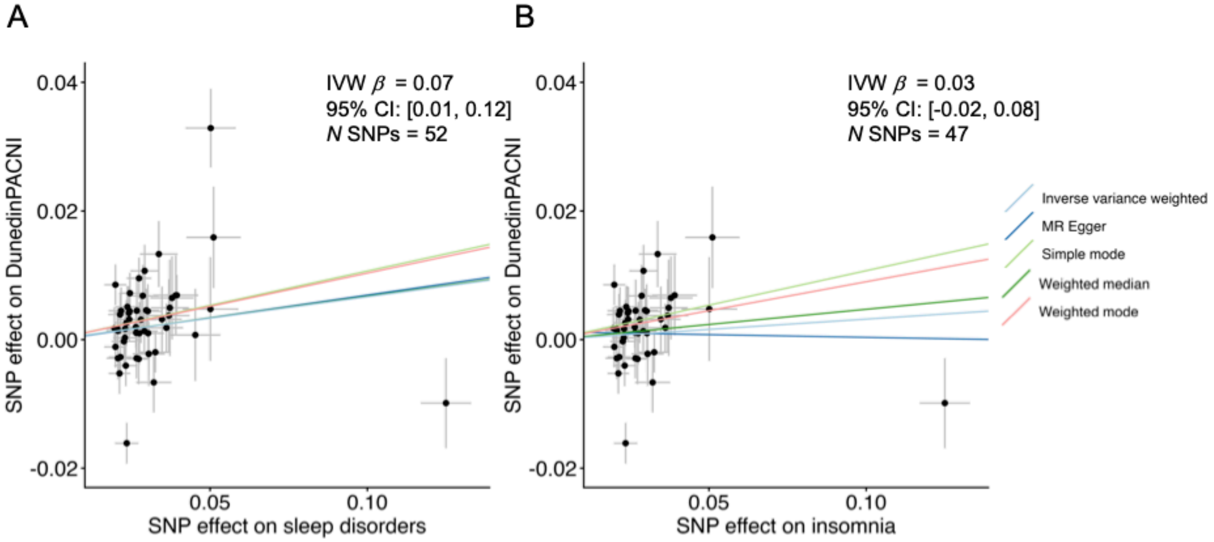

**Testing the effect of SNPs associated with confounding phenotypes on the causal influence of sleep disorders on DunedinPACNI. A.** Original model using all 52 instrumental SNPs from the FinnGen and MVP sleep disorder GWAS. **B.** Model after removing potentially confounded SNPs.

Collectively, these analyses suggest a casual association between sleep disorders and faster aging, however, some sensitivity tests indicate this finding may be influenced by horizontal pleiotropy. Therefore, we attempted to replicate this analysis using a GWAS of a more restricted sleep disorder phenotype in FinnGen.

2. *Effect of sleep disorders on DunedinPACNI with instrumental SNPs from the FinnGen non-organic sleep disorder GWAS*

Next, we tested the effects of sleep disorders on DunedinPACNI using genetic instruments identified from the GWAS of non-organic sleep disorders in FinnGen. We found evidence for a causal effect of non-organic sleep disorders on DunedinPACNI from inverse-variance weighted meta-analytic testing ($\beta$= 0.04, $p$=0.04, 95% CI: 0.00-0.07). Additional tests were in the same direction and of a similar magnitude.

**MR tests of the effect of non-organic sleep disorders on DunedinPACNI using instrumental SNPs from the FinnGen GWAS.**

| Exposure | Exposure instrument | Outcome | Outcome instrument | Method | *N* SNP | *b* | *p* |
|---|---|---|---|---|---|---|---|
| Non-organic sleep disorders | FinnGen GWAS | DunedinPACNI | UKB imaging GWAS | MR Egger | 7 | 0.09 | 0.15 |
| | | | | Weighted median | 7 | 0.05 | 0.02 |
| | | | | Inverse variance weighted | 7 | 0.04 | 0.04 |
| | | | | Simple mode | 7 | 0.08 | 0.11 |
| | | | | Weighted mode | 7 | 0.08 | 0.13 |

We did not observe heterogeneity between these instrumental SNPs (Q = 5.57, $p$ = 0.35), nor did we observe evidence of directional horizontal pleiotropy (MR-Egger intercept = -0.006, $p$ = 0.35). We visualized effects of each SNP using a forest plot. To further assess the robustness of our findings, we performed a leave-one-out analysis for each instrumental SNP. We observed consistent effects across leave-one-out tests, suggesting that this result was not driven by any specific SNP.

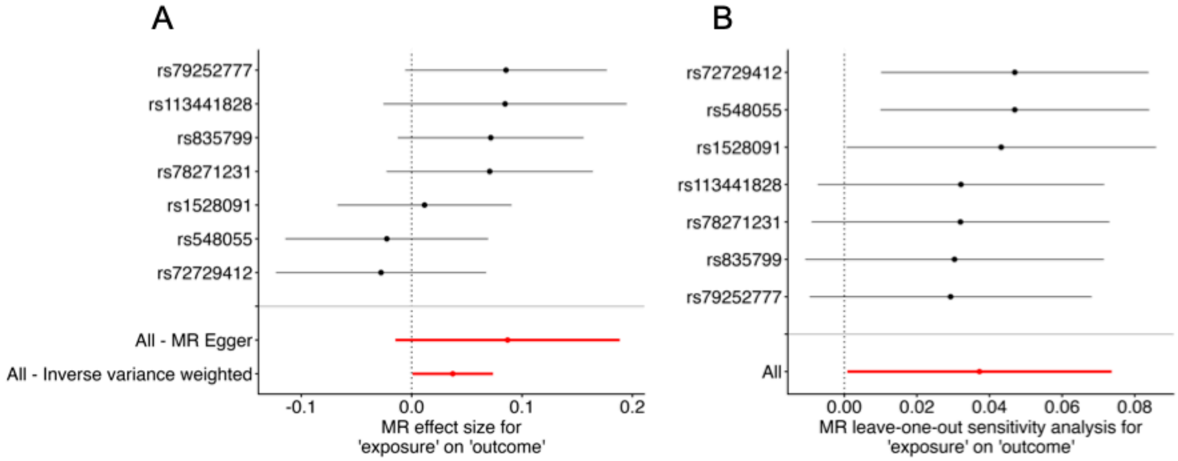

**Sensitivity tests of FinnGen non-organic sleep disorder GWAS meta-analysis MR analyses. A.** Forest plot of SNP MR effect of sleep disorders on DunedinPACNI. **B.** Forest plot of leave-one-out effects of sleep disorders on DunedinPACNI. Each row represents the row when that SNP is left out.

We formally tested for horizontal pleiotropy effects in this analysis using MR-PRESSO (Verbanck et al. 2018). The MR-PRESSO global test did not identify evidence of horizontal pleiotropy ($p$ = 0.39).

Finally, we again performed a biologically grounded sensitivity test for horizontal pleiotropy by searching for other genome-wide significant associations for instrumental SNPs using the *gwasrapidd* R package (Magno and Maia 2020). We did not observe any other genome-wide significant associations of the SNPs used as instrumental variables in this analysis.

Collectively, these results present evidence for a causal influence of sleep disorders on faster aging. We observed statistically significant effects using inverse-variance weighted tests with sets of genetic instruments for sleep disorders using more and less relaxed categorizations. Further, all alternative testing approaches yielded estimates in the same direction and of a similar magnitude. Finally, formal correction for horizontally pleiotropic SNPs strengthened evidence for a causal influence if sleep disorders on DunedinPACNI.

3. *Effect of DunedinPACNI on sleep disorders with instrumental SNPs from the UK Biobank DunedinPACNI GWAS*

Finally, we tested for evidence of a causal effect of DunedinPACNI on sleep disorders using the genetic instruments identified in the DunedinPACNI GWAS described above. We identified 18 instrumental SNPs for DunedinPACNI from the GWAS described above, shown in **Supplemental Table X**. We first tested the influence of DunedinPACNI on sleep disorders using the summary statistics from the GWAS meta-analysis of sleep disorders performed in FinnGen and the MVP. This analysis showed trend-level evidence of a causal effect (inverse-variance weighted $\beta$=0.17, $p$ = 0.08, 95% CI = [-0.02-0.35], for other methods see table below.

**MR tests of the effect of DunedinPACNI on sleep disorders using instrumental SNPs from the UK Biobank GWAS.**

| Exposure | Exposure instrument | Outcome | Outcome instrument | Method | *N* SNP | *b* | *p* |
|---|---|---|---|---|---|---|---|
| | | | | MR Egger | 18 | 0.76 | 0.14 |
| | | | | Weighted median | 18 | 0.15 | 0.04 |
| DunedinPACNI | UKB imaging GWAS | Sleep disorders | FinnGen + MVP GWAS | Inverse variance weighted | 18 | 0.17 | 0.08 |
| | | | | Simple mode | 18 | 0.17 | 0.17 |
| | | | | Weighted mode | 18 | 0.18 | 0.18 |

We observed heterogeneity between these instrumental SNPs (Q = 66.92, $p < 0.001$), though we did not observe evidence of directional horizontal pleiotropy (MR-Egger intercept = -0.01, $p = 0.23$). Leave-one-out analysis revealed evidence that the effects of *rs6546032* and *rs895932* may have driven the results toward the null.

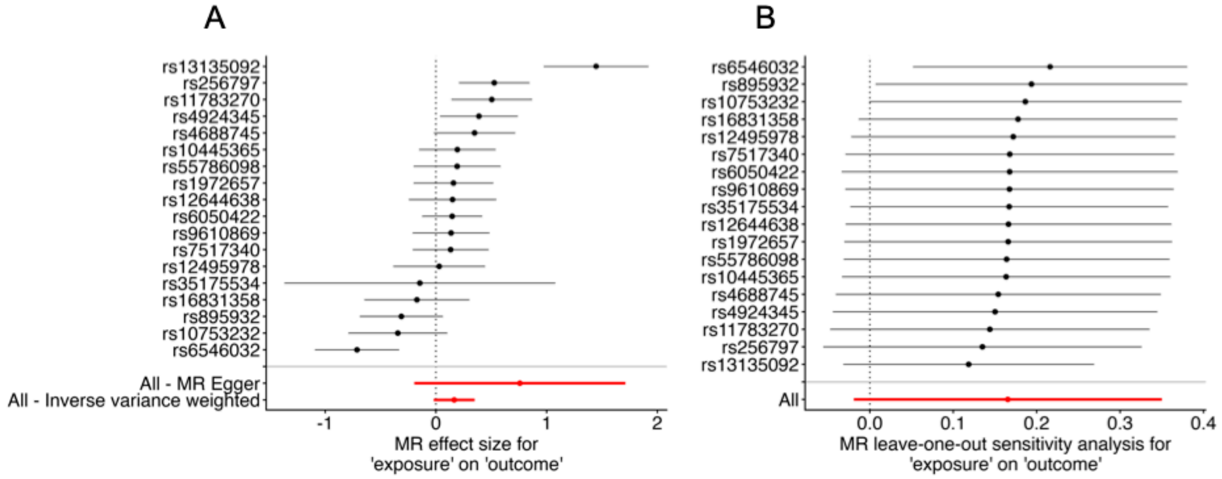

**Sensitivity tests of DunedinPACNI GWAS MR analyses using the FinnGen and MVP GWAS meta-analysis. A.** Forest plot of SNP effects of DunedinPACNI on sleep disorders. **B.** Forest plot of leave-one-out effects of DunedinPACNI on sleep disorders. Each row represents the row when that SNP is left out.

We formally tested for and corrected for horizontal pleiotropy effects in this analysis using MR-PRESSO (Verbanck et al. 2018). The MR-PRESSO global test identified evidence of horizontal pleiotropy ($p < 0.001$) driven by *rs6546032, rs13135092.* The outlier-corrected inverse variance weighted causal estimate was highly similar to the original estimate (Outlier-corrected: $\beta$=0.17, $p$=0.006; original: $\beta$=0.17, $p$=0.08), suggesting that horizontally pleiotropic SNPs may have drawn the effect towards the null.

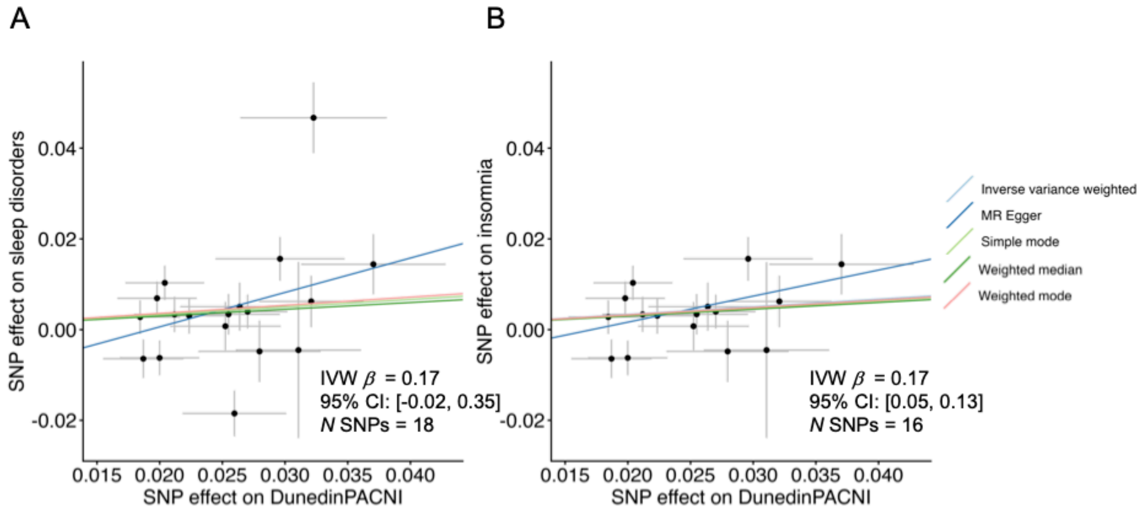

**MR-PRESSO outlier-correction of the effect of DunedinPACNI on sleep disorders. A.** Original model using all 52 instrumental SNPs from the FinnGen and MVP sleep disorder GWAS. **B.** Model after using MR-PRESSO to remove outlying SNPs.

4. *Effect of DunedinPACNI on non-organic sleep disorders with instrumental SNPs from the UK Biobank DunedinPACNI GWAS*

We next tested the influence of DunedinPACNI on non-organic sleep disorders using the summary statistics from the GWAS of non-organic sleep disorders performed in FinnGen. This analysis did not show evidence of a causal effect (inverse-variance weighted $\beta$=0.04, $p$ = 0.89, 95% CI = [-0.56-0.64], for other methods see table below.

**MR tests of the effect of DunedinPACNI on sleep disorders using instrumental SNPs from the UK Biobank GWAS.**

| Exposure | Exposure instrument | Outcome | Outcome instrument | Method | *N* SNP | *b* | *p* |
|---|---|---|---|---|---|---|---|
| DunedinPACNI | UKB imaging GWAS | Non-organic sleep disorders | FinnGen GWAS | MR Egger | 16 | 0.14 | 0.94 |
| | | | | Weighted median | 16 | 0.57 | 0.08 |
| | | | | Inverse variance weighted | 16 | 0.04 | 0.89 |
| | | | | Simple mode | 16 | 0.85 | 0.47 |
| | | | | Weighted mode | 16 | 0.80 | 0.50 |

We observed heterogeneity between these instrumental SNPs (MR-Egger Q = 28.76, $p$ = 0.01), though we did not observe evidence of directional horizontal pleiotropy (MR-Egger intercept = -0.002, $p$ = 0.95). Leave-one-out analysis did not reveal evidence that any individual SNP drove the results toward the null.

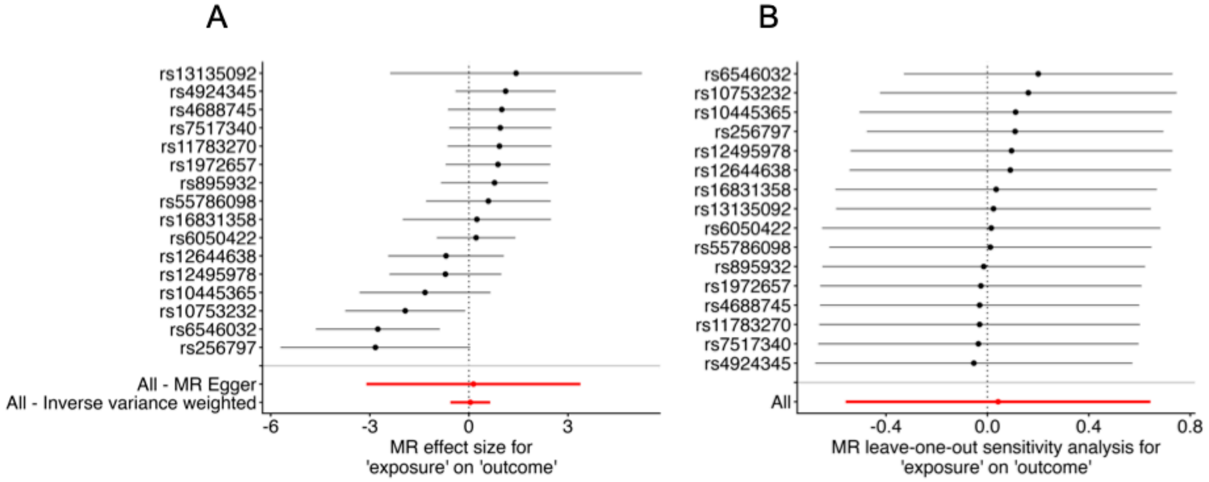

**Sensitivity tests of DunedinPACNI GWAS MR analyses using the FinnGen GWAS. A.** Forest plot of SNP effects of DunedinPACNI on non-organic sleep disorders. **B.** Forest plot of leave-one-out effects of DunedinPACNI on non-organic sleep disorders. Each row represents the row when that SNP is left out.

We formally tested for and corrected for horizontal pleiotropy effects in this analysis using MR-PRESSO (Verbanck et al. 2018). The MR-PRESSO global test identified evidence of horizontal pleiotropy ($p$ = 0.02) driven by *rs6546032.* The outlier-corrected inverse variance weighted causal estimate was much stronger than the original estimate, though was still null (Outlier-corrected: $\beta$=0.20, $p$=0.45; original: $\beta$=0.04, $p$=0.89), providing little evidence of a casual effect and suggesting instability of the model to outlying SNPs.

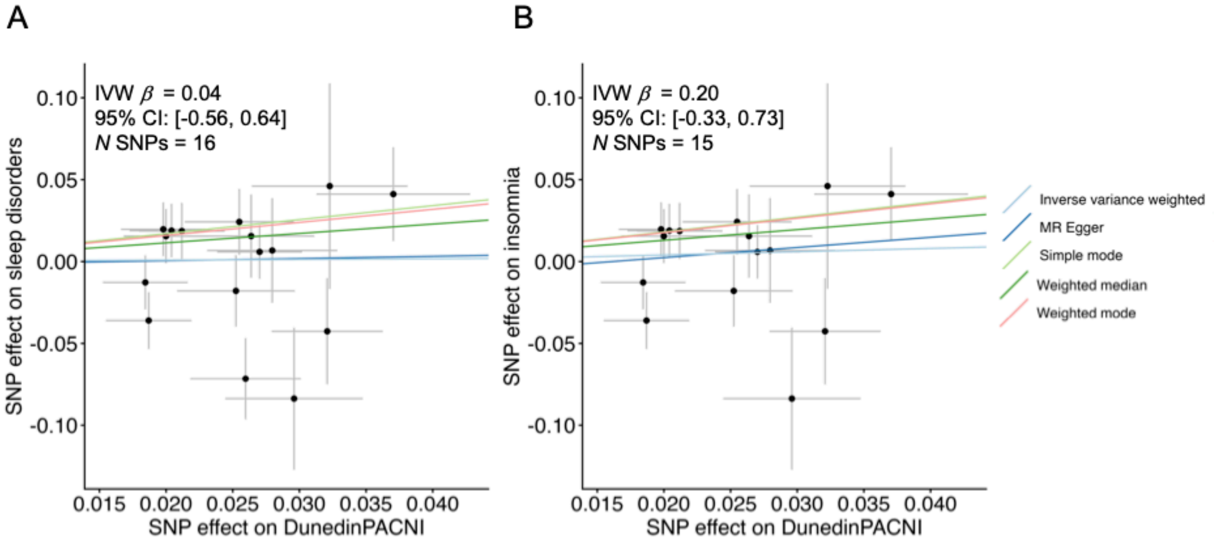

**MR-PRESSO outlier-correction of the effect of DunedinPACNI on non-organic sleep disorders. A.** Original model using all 16 instrumental SNPs from the FinnGen non-organic sleep disorder GWAS. **B.** Model after using MR-PRESSO to remove the outlying SNP.

Collectively, these analyses find limited evidence for a causal influence of DunedinPACNI on sleep disorders. Although we observed a trend-level effect of DunedinPACNI on sleep disorders using the FinnGen and MVP GWAS meta-analytic summary statistics, this result was not replicated when using the GWAS of non-organic sleep disorders in FinnGen. Overall, this does not present strong evidence of a causal impact of accelerated aging on sleep disorders.

**Supplemental Table S1. Effects of controlling for chronic health conditions on the association between PSQI scores and DunedinPACE in MIDUS**

| Model | *N* | beta | *p* | 95% CI |
|---|---|---|---|---|
| No control | 983 | 0.14 | < .001*** | [0.08, 0.20] |
| Controlling for disease | 983 | 0.10 | .001** | [0.04, 0.16] |

Abbreviations: CI = confidence interval, MIDUS = Midlife Development in the United States.

**Supplemental Table S2. Effects of controlling for Charlson Comorbidity Index on the associations between sleep and DunedinPACNI in the UK Biobank**

| Model | *N* | Model version | beta | *p* | 95% CI |
|---|---|---|---|---|---|
| Short vs. Average sleep | 50,743 | Original model | 0.05 | < .001*** | [0.04, 0.07] |
| | 50,743 | CCI control | 0.05 | < .001*** | [0.03, 0.07] |
| Long vs. Average sleep | 38,559 | Original model | 0.27 | < .001*** | [0.21, 0.33] |
| | 38,559 | CCI control | 0.25 | < .001*** | [0.19, 0.31] |
| Sometimes vs. Never insomnia | 34,020 | Original model | -0.00 | .695 | [-0.23, 0.02] |
| | 34,020 | CCI control | -0.01 | .569 | [-0.25, 0.14] |
| Usually vs. Never insomnia | 28,204 | Original model | 0.03 | .002 | [0.01, 0.05] |
| | 28,204 | CCI control | 0.02 | .020 | [0.00, 0.04] |

**Supplemental Table S3. Effects of controlling for cognitive status on the associations between informant-reported sleep and aging in ADNI**

| | | *N* | | Original model | | | Controlling for cognitive status | | |
|---|---|---|---|---|---|---|---|---|---|
| **Measure** | **Comparison** | **Individuals** | **Observations** | ***β*** | ***p*** | **95% CI** | ***β*** | ***p*** | **95% CI** |
| DunedinPACNI | Mild vs. None | 744 | 1,966 | 0.26 | .002** | [0.10, 0.43] | 0.18 | .022* | [0.03, 0.33] |
| | Moderate vs. None | 712 | 1,826 | 0.35 | .002** | [0.13, 0.56] | 0.18 | .096 | [-0.03, 0.38] |
| | Severe vs. None | 695 | 1,749 | 0.87 | .001*** | [0.37, 1.37] | 0.59 | .008** | [0.15, 1.03] |
| DunedinPACE | Mild vs. None | 490 | 731 | 0.22 | .068 | [-0.02, 0.46] | 0.19 | .121 | [-0.05, 0.43] |
| | Moderate vs. None | 481 | 687 | 0.02 | .849 | [-0.20, 0.24] | -0.04 | .743 | [-0.26, 0.19] |
| | Severe vs. None | 449 | 643 | 0.55 | .020* | [0.09, 1.02] | 0.50 | .038* | [0.03, 0.96] |

Abbreviations: ADNI = Alzheimer’s Disease Neuroimaging Initiative, CI = Confidence interval.

**Supplemental Table S4. MIDUS sample demographic information**

| Sample | *N* | Mean age | SD age | Sex (% female) | Mean PSQI | SD PSQI |
|---|---|---|---|---|---|---|
| Original | 1,207 | 53.97 | 12.58 | 56 | 6.14 | 3.50 |
| Chronic disease | 983 | 54.60 | 12.81 | 54 | 5.79 | 3.29 |

*Note.* Abbreviations: MIDUS = Midlife development in the United States, PSQI = Pittsburgh Sleep Quality Index, SD = standard deviation

**Supplemental Table S5. UK Biobank sample demographic information**

| Sample | *N* | Mean age | SD age | Sex (% female) | Mean sleep duration | SD sleep duration |
|---|---|---|---|---|---|---|
| Sleep duration | 59,844 | 65.01 | 7.74 | 53 | 7.15 | 1.06 |
| Insomnia | 59,970 | 65.01 | 7.74 | 53 | 7.15 | 1.06 |
| Sleep duration and CCI | 51,480 | 65.26 | 7.77 | 54 | 7.14 | 1.07 |
| Insomnia and CCI | 51,602 | 65.26 | 7.77 | 54 | 7.14 | 1.07 |

*Note.* Abbreviations: CCI = Charlson Comorbidity Index, SD = standard deviation

**Supplemental Table S6. UK Biobank GWAS sample demographic information**

| Sample | *N* | Mean age | SD age | Min age | Max age | Sex (% female) | N never/rarely insomnia | N sometimes insomnia | N usually insomnia | Mean DunedinPACNI | SD DunedinPACNI | Min DunedinPACNI | Max DunedinPACNI |
|---|---|---|---|---|---|---|---|---|---|---|---|---|---|
| DunedinPACNI | 244,436 | 57.15 | 8.03 | 39 | 72 | 53.2 | - | - | - | 1.20 | 0.70 | -1.50 | 4.75 |
| Insomnia | 65,868 | 66.07 | 7.88 | 45 | 87 | 53.6 | 57,570 | 116,638 | 70,228 | - | - | - | - |

Age presented in years. Abbreviations: Max = maximum, Min = minimum, SD = standard deviation

**Supplemental Table S7. ICD10 codes used to estimate Charlson Comorbidity Index in UK Biobank**

| Diagnosis | ICD10 three character | ICD10 fourth character | Charlson weight |
|---|---|---|---|
| Congestive heart failure | I09 | 9 | 1 |
| Congestive heart failure | I11 | 0 | 1 |
| Congestive heart failure | I13 | 0 | 1 |
| Congestive heart failure | I13 | 2 | 1 |
| Congestive heart failure | I25 | 5 | 1 |
| Congestive heart failure | I42 | 0 | 1 |
| Congestive heart failure | I42 | 5 | 1 |
| Congestive heart failure | I42 | 6 | 1 |
| Congestive heart failure | I42 | 7 | 1 |
| Congestive heart failure | I42 | 8 | 1 |
| Congestive heart failure | I42 | 9 | 1 |
| Congestive heart failure | I43 | | 1 |
| Congestive heart failure | I50 | | 1 |
| Congestive heart failure | P29 | 0 | 1 |
| Cardiac arrhythmias | I44 | 1 | 1 |
| Cardiac arrhythmias | I44 | 2 | 1 |
| Cardiac arrhythmias | I44 | 3 | 1 |
| Cardiac arrhythmias | I45 | 6 | 1 |
| Cardiac arrhythmias | I45 | 9 | 1 |
| Cardiac arrhythmias | I47 | | 1 |
| Cardiac arrhythmias | I48 | | 1 |
| Cardiac arrhythmias | I49 | | 1 |
| Cardiac arrhythmias | R00 | 0 | 1 |
| Cardiac arrhythmias | R00 | 1 | 1 |
| Cardiac arrhythmias | R00 | 8 | 1 |
| Cardiac arrhythmias | T82 | 1 | 1 |

| | | | |
|---|---|---|---|
| Cardiac arrhythmias | Z45 | 0 | 1 |
| Cardiac arrhythmias | Z95 | 0 | 1 |
| Vascular disease | A52 | 0 | 1 |
| Vascular disease | I05 | | 1 |
| Vascular disease | I06 | | 1 |
| Vascular disease | I07 | | 1 |
| Vascular disease | I08 | | 1 |
| Vascular disease | I09 | 1 | 1 |
| Vascular disease | I09 | 8 | 1 |
| Vascular disease | I34 | | 1 |
| Vascular disease | I35 | | 1 |
| Vascular disease | I36 | | 1 |
| Vascular disease | I37 | | 1 |
| Vascular disease | I38 | | 1 |
| Vascular disease | I39 | | 1 |
| Vascular disease | Q23 | 0 | 1 |
| Vascular disease | Q23 | 1 | 1 |
| Vascular disease | Q23 | 2 | 1 |
| Vascular disease | Q23 | 3 | 1 |
| Vascular disease | Z95 | 2 | 1 |
| Vascular disease | Z95 | 3 | 1 |
| Vascular disease | Z95 | 4 | 1 |
| Pulmonary circulation disorders | I26 | | 1 |
| Pulmonary circulation disorders | I27 | | 1 |
| Pulmonary circulation disorders | I28 | 0 | 1 |
| Pulmonary circulation disorders | I28 | 8 | 1 |
| Pulmonary circulation disorders | I28 | 9 | 1 |
| Peripheral vascular disorders | I70 | | 1 |
| Peripheral vascular disorders | I71 | | 1 |

| | | | |
|---|---|---|---|
| Peripheral vascular disorders | I73 | 1 | 1 |
| Peripheral vascular disorders | I73 | 8 | 1 |
| Peripheral vascular disorders | I73 | 9 | 1 |
| Peripheral vascular disorders | I77 | 1 | 1 |
| Peripheral vascular disorders | I79 | 0 | 1 |
| Peripheral vascular disorders | I79 | 2 | 1 |
| Peripheral vascular disorders | K55 | 1 | 1 |
| Peripheral vascular disorders | K55 | 8 | 1 |
| Peripheral vascular disorders | Z95 | 8 | 1 |
| Peripheral vascular disorders | Z95 | 9 | 1 |
| Hypertension, uncomplicated | I10 | | 1 |
| Hypertension, complicated | I11 | | 1 |
| Hypertension, complicated | I12 | | 1 |
| Hypertension, complicated | I13 | | 1 |
| Hypertension, complicated | I15 | | 1 |
| Paralysis | G04 | 1 | 2 |
| Paralysis | G11 | 4 | 2 |
| Paralysis | G80 | 1 | 2 |
| Paralysis | G80 | 2 | 2 |
| Paralysis | G81 | | 2 |
| Paralysis | G82 | | 2 |
| Paralysis | G83 | 0 | 2 |
| Paralysis | G83 | 1 | 2 |
| Paralysis | G83 | 2 | 2 |
| Paralysis | G83 | 3 | 2 |
| Paralysis | G83 | 4 | 2 |
| Paralysis | G83 | 9 | 2 |
| Chronic pulmonary disease | I27 | 8 | 1 |
| Chronic pulmonary disease | I27 | 9 | 1 |

| | | | |
|---|---|---|---|
| Chronic pulmonary disease | J40 | | 1 |
| Chronic pulmonary disease | J41 | | 1 |
| Chronic pulmonary disease | J42 | | 1 |
| Chronic pulmonary disease | J43 | | 1 |
| Chronic pulmonary disease | J44 | | 1 |
| Chronic pulmonary disease | J45 | | 1 |
| Chronic pulmonary disease | J46 | | 1 |
| Chronic pulmonary disease | J47 | | 1 |
| Chronic pulmonary disease | J60 | | 1 |
| Chronic pulmonary disease | J61 | | 1 |
| Chronic pulmonary disease | J62 | | 1 |
| Chronic pulmonary disease | J63 | | 1 |
| Chronic pulmonary disease | J64 | | 1 |
| Chronic pulmonary disease | J65 | | 1 |
| Chronic pulmonary disease | J66 | | 1 |
| Chronic pulmonary disease | J67 | | 1 |
| Chronic pulmonary disease | J68 | 4 | 1 |
| Chronic pulmonary disease | J70 | 1 | 1 |
| Chronic pulmonary disease | J70 | 3 | 1 |
| Diabetes, uncomplicated | E10 | 0 | 1 |
| Diabetes, uncomplicated | E10 | 1 | 1 |
| Diabetes, uncomplicated | E10 | 9 | 1 |
| Diabetes, uncomplicated | E11 | 0 | 1 |
| Diabetes, uncomplicated | E11 | 1 | 1 |
| Diabetes, uncomplicated | E11 | 9 | 1 |
| Diabetes, uncomplicated | E12 | 0 | 1 |
| Diabetes, uncomplicated | E12 | 1 | 1 |
| Diabetes, uncomplicated | E12 | 9 | 1 |
| Diabetes, uncomplicated | E13 | 0 | 1 |

| | | | |
|---|---|---|---|
| Diabetes, uncomplicated | E13 | 1 | 1 |
| Diabetes, uncomplicated | E13 | 9 | 1 |
| Diabetes, uncomplicated | E14 | 0 | 1 |
| Diabetes, uncomplicated | E14 | 1 | 1 |
| Diabetes, uncomplicated | E14 | 9 | 1 |
| Diabetes, complicated | E10 | 2 | 2 |
| Diabetes, complicated | E10 | 3 | 2 |
| Diabetes, complicated | E10 | 4 | 2 |
| Diabetes, complicated | E10 | 5 | 2 |
| Diabetes, complicated | E10 | 6 | 2 |
| Diabetes, complicated | E10 | 7 | 2 |
| Diabetes, complicated | E10 | 8 | 2 |
| Diabetes, complicated | E11 | 2 | 2 |
| Diabetes, complicated | E11 | 3 | 2 |
| Diabetes, complicated | E11 | 4 | 2 |
| Diabetes, complicated | E11 | 5 | 2 |
| Diabetes, complicated | E11 | 6 | 2 |
| Diabetes, complicated | E11 | 7 | 2 |
| Diabetes, complicated | E11 | 8 | 2 |
| Diabetes, complicated | E12 | 2 | 2 |
| Diabetes, complicated | E12 | 3 | 2 |
| Diabetes, complicated | E12 | 4 | 2 |
| Diabetes, complicated | E12 | 5 | 2 |
| Diabetes, complicated | E12 | 6 | 2 |
| Diabetes, complicated | E12 | 7 | 2 |
| Diabetes, complicated | E12 | 8 | 2 |
| Diabetes, complicated | E13 | 2 | 2 |
| Diabetes, complicated | E13 | 3 | 2 |
| Diabetes, complicated | E13 | 4 | 2 |

| | | | |
|---|---|---|---|
| Diabetes, complicated | E13 | 5 | 2 |
| Diabetes, complicated | E13 | 6 | 2 |
| Diabetes, complicated | E13 | 7 | 2 |
| Diabetes, complicated | E13 | 8 | 2 |
| Diabetes, complicated | E14 | 2 | 2 |
| Diabetes, complicated | E14 | 3 | 2 |
| Diabetes, complicated | E14 | 4 | 2 |
| Diabetes, complicated | E14 | 5 | 2 |
| Diabetes, complicated | E14 | 6 | 2 |
| Diabetes, complicated | E14 | 7 | 2 |
| Diabetes, complicated | E14 | 8 | 2 |
| Renal failure | I12 | 0 | 2 |
| Renal failure | I13 | 1 | 2 |
| Renal failure | N18 | | 2 |
| Renal failure | N19 | | 2 |
| Renal failure | N25 | 0 | 2 |
| Renal failure | Z49 | 0 | 2 |
| Renal failure | Z49 | 1 | 2 |
| Renal failure | Z49 | 2 | 2 |
| Renal failure | Z94 | 0 | 2 |
| Renal failure | Z99 | 2 | 2 |
| Liver disease | B18 | | 3 |
| Liver disease | I85 | | 3 |
| Liver disease | I86 | 4 | 3 |
| Liver disease | I98 | 2 | 3 |
| Liver disease | K70 | | 3 |
| Liver disease | K71 | 1 | 3 |
| Liver disease | K71 | 3 | 3 |
| Liver disease | K71 | 4 | 3 |

| | | | |
|---|---|---|---|
| Liver disease | K71 | 5 | 3 |
| Liver disease | K71 | 7 | 3 |
| Liver disease | K72 | | 3 |
| Liver disease | K73 | | 3 |
| Liver disease | K74 | | 3 |
| Liver disease | K76 | 0 | 3 |
| Liver disease | K76 | 2 | 3 |
| Liver disease | K76 | 3 | 3 |
| Liver disease | K76 | 4 | 3 |
| Liver disease | K76 | 5 | 3 |
| Liver disease | K76 | 6 | 3 |
| Liver disease | K76 | 7 | 3 |
| Liver disease | K76 | 8 | 3 |
| Liver disease | K76 | 9 | 3 |
| Liver disease | Z94 | 4 | 3 |
| Peptic ulcer disease, excluding bleeding | K25 | 7 | 1 |
| Peptic ulcer disease, excluding bleeding | K25 | 9 | 1 |
| Peptic ulcer disease, excluding bleeding | K26 | 7 | 1 |
| Peptic ulcer disease, excluding bleeding | K26 | 9 | 1 |
| Peptic ulcer disease, excluding bleeding | K27 | 7 | 1 |
| Peptic ulcer disease, excluding bleeding | K27 | 9 | 1 |
| Peptic ulcer disease, excluding bleeding | K28 | 7 | 1 |
| Peptic ulcer disease, excluding bleeding | K28 | 9 | 1 |
| AIDS/HIV | B20 | | 6 |
| AIDS/HIV | B21 | | 6 |
| AIDS/HIV | B22 | | 6 |
| AIDS/HIV | B24 | | 6 |
| Lymphoma | C81 | | 2 |
| Lymphoma | C82 | | 2 |

| | | | |
|---|---|---|---|
| Lymphoma | C83 | | 2 |
| Lymphoma | C84 | | 2 |
| Lymphoma | C85 | | 2 |
| Lymphoma | C88 | | 2 |
| Lymphoma | C96 | | 2 |
| Lymphoma | C90 | 0 | 2 |
| Lymphoma | C90 | 2 | 2 |
| Metastatic cancer | C77 | | 6 |
| Metastatic cancer | C78 | | 6 |
| Metastatic cancer | C79 | | 6 |
| Metastatic cancer | C80 | | 6 |
| Solid tumour without metastasis | C00 | | 2 |
| Solid tumour without metastasis | C01 | | 2 |
| Solid tumour without metastasis | C02 | | 2 |
| Solid tumour without metastasis | C03 | | 2 |
| Solid tumour without metastasis | C04 | | 2 |
| Solid tumour without metastasis | C05 | | 2 |
| Solid tumour without metastasis | C06 | | 2 |
| Solid tumour without metastasis | C07 | | 2 |
| Solid tumour without metastasis | C08 | | 2 |
| Solid tumour without metastasis | C09 | | 2 |
| Solid tumour without metastasis | C10 | | 2 |
| Solid tumour without metastasis | C11 | | 2 |
| Solid tumour without metastasis | C12 | | 2 |
| Solid tumour without metastasis | C13 | | 2 |
| Solid tumour without metastasis | C14 | | 2 |
| Solid tumour without metastasis | C15 | | 2 |
| Solid tumour without metastasis | C16 | | 2 |
| Solid tumour without metastasis | C17 | | 2 |

| | | |
|---|---|---|
| Solid tumour without metastasis | C18 | 2 |
| Solid tumour without metastasis | C19 | 2 |
| Solid tumour without metastasis | C20 | 2 |
| Solid tumour without metastasis | C21 | 2 |
| Solid tumour without metastasis | C22 | 2 |
| Solid tumour without metastasis | C23 | 2 |
| Solid tumour without metastasis | C24 | 2 |
| Solid tumour without metastasis | C25 | 2 |
| Solid tumour without metastasis | C26 | 2 |
| Solid tumour without metastasis | C30 | 2 |
| Solid tumour without metastasis | C31 | 2 |
| Solid tumour without metastasis | C32 | 2 |
| Solid tumour without metastasis | C33 | 2 |
| Solid tumour without metastasis | C34 | 2 |
| Solid tumour without metastasis | C37 | 2 |
| Solid tumour without metastasis | C38 | 2 |
| Solid tumour without metastasis | C39 | 2 |
| Solid tumour without metastasis | C40 | 2 |
| Solid tumour without metastasis | C41 | 2 |
| Solid tumour without metastasis | C43 | 2 |
| Solid tumour without metastasis | C45 | 2 |
| Solid tumour without metastasis | C46 | 2 |
| Solid tumour without metastasis | C47 | 2 |
| Solid tumour without metastasis | C48 | 2 |
| Solid tumour without metastasis | C49 | 2 |
| Solid tumour without metastasis | C50 | 2 |
| Solid tumour without metastasis | C51 | 2 |
| Solid tumour without metastasis | C52 | 2 |
| Solid tumour without metastasis | C53 | 2 |

| | | | |
|---|---|---|---|
| Solid tumour without metastasis | C54 | | 2 |
| Solid tumour without metastasis | C55 | | 2 |
| Solid tumour without metastasis | C56 | | 2 |
| Solid tumour without metastasis | C57 | | 2 |
| Solid tumour without metastasis | C58 | | 2 |
| Solid tumour without metastasis | C60 | | 2 |
| Solid tumour without metastasis | C61 | | 2 |
| Solid tumour without metastasis | C62 | | 2 |
| Solid tumour without metastasis | C63 | | 2 |
| Solid tumour without metastasis | C64 | | 2 |
| Solid tumour without metastasis | C65 | | 2 |
| Solid tumour without metastasis | C66 | | 2 |
| Solid tumour without metastasis | C67 | | 2 |
| Solid tumour without metastasis | C68 | | 2 |
| Solid tumour without metastasis | C69 | | 2 |
| Solid tumour without metastasis | C70 | | 2 |
| Solid tumour without metastasis | C71 | | 2 |
| Solid tumour without metastasis | C72 | | 2 |
| Solid tumour without metastasis | C73 | | 2 |
| Solid tumour without metastasis | C74 | | 2 |
| Solid tumour without metastasis | C75 | | 2 |
| Solid tumour without metastasis | C76 | | 2 |
| Solid tumour without metastasis | C97 | | 2 |
| Rheumatoid arthritis/collagen vascular diseases | L94 | 0 | 1 |
| Rheumatoid arthritis/collagen vascular diseases | L94 | 1 | 1 |
| Rheumatoid arthritis/collagen vascular diseases | L94 | 3 | 1 |
| Rheumatoid arthritis/collagen vascular diseases | M05 | | 1 |
| Rheumatoid arthritis/collagen vascular diseases | M06 | | 1 |
| Rheumatoid arthritis/collagen vascular diseases | M08 | | 1 |

| | | | |
|---|---|---|---|
| Rheumatoid arthritis/collagen vascular diseases | M12 | 0 | 1 |
| Rheumatoid arthritis/collagen vascular diseases | M12 | 3 | 1 |
| Rheumatoid arthritis/collagen vascular diseases | M30 | | 1 |
| Rheumatoid arthritis/collagen vascular diseases | M31 | 0 | 1 |
| Rheumatoid arthritis/collagen vascular diseases | M31 | 1 | 1 |
| Rheumatoid arthritis/collagen vascular diseases | M31 | 2 | 1 |
| Rheumatoid arthritis/collagen vascular diseases | M31 | 3 | 1 |
| Rheumatoid arthritis/collagen vascular diseases | M32 | | 1 |
| Rheumatoid arthritis/collagen vascular diseases | M33 | | 1 |
| Rheumatoid arthritis/collagen vascular diseases | M34 | | 1 |
| Rheumatoid arthritis/collagen vascular diseases | M35 | | 1 |
| Rheumatoid arthritis/collagen vascular diseases | M45 | | 1 |
| Rheumatoid arthritis/collagen vascular diseases | M46 | 1 | 1 |
| Rheumatoid arthritis/collagen vascular diseases | M46 | 8 | 1 |
| Rheumatoid arthritis/collagen vascular diseases | M46 | 9 | 1 |
| Coagulopathy | D65 | | 1 |
| Coagulopathy | D66 | | 1 |
| Coagulopathy | D67 | | 1 |
| Coagulopathy | D68 | | 1 |
| Coagulopathy | D69 | 1 | 1 |
| Coagulopathy | D69 | 3 | 1 |
| Coagulopathy | D69 | 4 | 1 |
| Coagulopathy | D69 | 5 | 1 |
| Coagulopathy | D69 | 6 | 1 |
| Blood loss anaemia | D50 | 0 | 1 |
| Deficiency anaemia | D50 | 8 | 1 |
| Deficiency anaemia | D50 | 9 | 1 |
| Deficiency anaemia | D51 | | 1 |
| Deficiency anaemia | D52 | | 1 |

| | | |
|---|---|---|
| Deficiency anaemia | D53 | 1 |

Abbreviations: ICD = International Classification of Diseases

**Supplemental Table S8. ADNI sample demographic information**

| | *N* | | | | | | | |
|---|---|---|---|---|---|---|---|---|
| **Sample** | **Observations** | **Individuals** | **Mean age** | **SD age** | **Sex (% female)** | ***N* CN** | ***N* MCI** | ***N* Dementia** |
| MRI | 2,089 | 759 | 75.57 | 6.67 | 44 | 652 | 907 | 530 |
| DNAm | 809 | 532 | 75.40 | 7.72 | 44 | 244 | 409 | 156 |

Abbreviations: ADNI = Alzheimer's Disease Neuroimaging Initiative, DNAm = DNA methylation, CN = cognitively normal, MCI = mild cognitive impairment, MRI = magnetic resonance imaging, SD = standard deviation

**Supplemental Table S9. HCP sample demographic information**

| Sample | *N* | Mean age | SD age | Min age | Max age | Sex (% female) | Mean PSQI | SD PSQI |
|---|---|---|---|---|---|---|---|---|
| Overall | 1,113 | 28.80 | 3.70 | 22 | 37 | 54 | 4.80 | 2.76 |
| Monozygotic | 240 | 29.55 | 3.35 | 22 | 36 | 66 | 4.44 | 2.43 |
| Dizygotic | 238 | 29.05 | 3.57 | 22 | 36 | 56 | 4.61 | 2.82 |

*Note.* Abbreviations: PSQI = Pittsburgh Sleep Quality Index, SD = standard deviation

**Supplemental Table S10. E-Risk sample demographic information**

| Sample | *N* | N Families | Zygosity (% MZ) | Sex (% female) | Mean PSQI | SD PSQI |
|---|---|---|---|---|---|---|
| Original | 1,657 | 924 | 57 | 50 | 5.34 | 3.12 |
| Twin | 1,466 | 733 | 58 | 51 | 5.29 | 3.10 |

*Note.* Abbreviations: MZ = monozygotic, PSQI = Pittsburgh Sleep Quality Index, SD = standard deviation
